# Pulmonary inflammation in severe pneumonia is characterised by compartmentalised and mechanistically distinct sub-phenotypes

**DOI:** 10.1101/2024.09.02.24312971

**Authors:** M Jeffrey, J Bartholdson Scott, RJ White, E Higginson, M Maes, S Forrest, J Pereira-Dias, S Parmar, E Heasman-Hunt, MD Curran, P Polgarova, J Herre, EE Davenport, S Baker, G Dougan, V Navapurkar, A Conway Morris

## Abstract

Pneumonia is the leading infectious disease killer worldwide and commonly requires admission to critical care. Despite its prevalence, the underpinning biology of severe pneumonia remains incompletely understood. We performed multifaceted assessments of bronchoalveolar transcriptome, cytokines, microbiology, and clinical features to biologically dissect a cohort of patients with suspected severe pneumonia. Our data revealed three lung-restricted transcriptionally defined severe pneumonia endotypes (termed ‘Pneumotypes’ (Pn)). All three Pneumotypes had comparable clinical presentations and severity of respiratory failure but critically had divergent outcomes. Pn1, the most common, was characterised by low alveolar cytokines, expanded tolerogenic macrophages and epithelial damage. Pn3 was characterised by neutrophil-monocyte infiltration, IL-6-STAT3 activation and longer duration of mechanical ventilation. Pn2 displayed the fastest resolution, exhibiting a balanced immune response and epithelial-endothelial repair signatures. Our work has identified mechanistically distinct phenotypes in the lungs of patients with suspected pneumonia and acute lung injury, providing new targets for personalised therapy.

## Introduction

Pneumonia is the commonest infectious cause of death worldwide, responsible for an estimated 2.5 million deaths per year[1] and second only to diarrhoeal disease as a cause of sepsis[2]. Severe pneumonia contributes to a large proportion of the patient burden in intensive care units, accounting for 60% of all infections managed in this setting[3]. Pneumonia is also the most common trigger for acute respiratory distress syndrome (ARDS)[4], the development of which is associated with an increase in morbidity and mortality[5].

Despite the considerable burden of pneumonia, the syndrome is incompletely understood and diagnosis is difficult. There is limited overlap between the clinical-radiological syndrome used for diagnosis and histopathologically confirmed pneumonia[6]. Distinguishing infection from sterile mimics remains challenging[7, 8]. Blood-based biomarkers have poor diagnostic performance[9], leading to the investigation of lung sampling to identify compartmentalised inflammation. Whilst alveolar cytokines, notably interleukin 1 beta (IL-1β) and CXCL-8, have demonstrated excellent sensitivity, they have poor specificity[8] and failed to change antimicrobial prescribing in clinical trials[10]. Alveolar neutrophil counts are also sensitive but non-specific for pneumonia of bacterial origin[11]. Although the mechanisms driving alveolar inflammation remain unclear, the low specificity seen with both these measures implies common pathways terminating a diverse range of upstream insults.

Decades of limited therapeutic advances in critical illness have led to attempts to move away from broad, clinically defined syndromes and towards pathophysiologically defined entities[12]. Conflicting results in trials of immunomodulatory therapies in pneumonia maintain that this is also true for this clinical syndrome[13, 14]. Peripheral blood phenotypes have been identified in sepsis arising from pneumonia[15] and ARDS[16]. However, whilst these approaches predict outcomes and may explain some of the heterogeneity in therapeutic trials, to date, such an approach has not been applied directly at the site of infection, i.e. the lungs. This is despite the well-established compartmentalisation of inflammatory responses[8, 17].

We examined a cohort of ventilated patients with clinical pneumonia syndrome and profiled immune responses in bronchoalveolar lavage and blood compartments. We aimed to identify unrecognised heterogeneity and understand the diverse disease processes that may be exploited to personalise future pneumonia therapy.

## Results

### Bronchoalveolar host gene transcription defines three sub-phenotypes in patients with suspected pneumonia

We recruited a cohort of 95 mechanically ventilated patients with clinically suspected pneumonia from a mixed medical-surgical intensive care unit (ICU)[18]. Eighty of these patients had sequenceable RNA from bronchoalveolar lavage. The onset of suspected pneumonia was a mixture of community and hospital-acquired. Overall, 34 (43%) had their pneumonia confirmed after expert consensus review, with bacteria being the most common aetiological agents (Table 1). In keeping with the syndrome of severe pneumonia, the patients had significantly impaired oxygenation, a high rate of acute respiratory distress syndrome (ARDS) and high acute physiology and chronic health evaluation II (APACHE II) scores (a measure of the severity of illness)(Table 1). Rates of immunosuppression were also high at 40%, although not dissimilar to previously published cohorts of severe pneumonia [19].

**Table 1.**
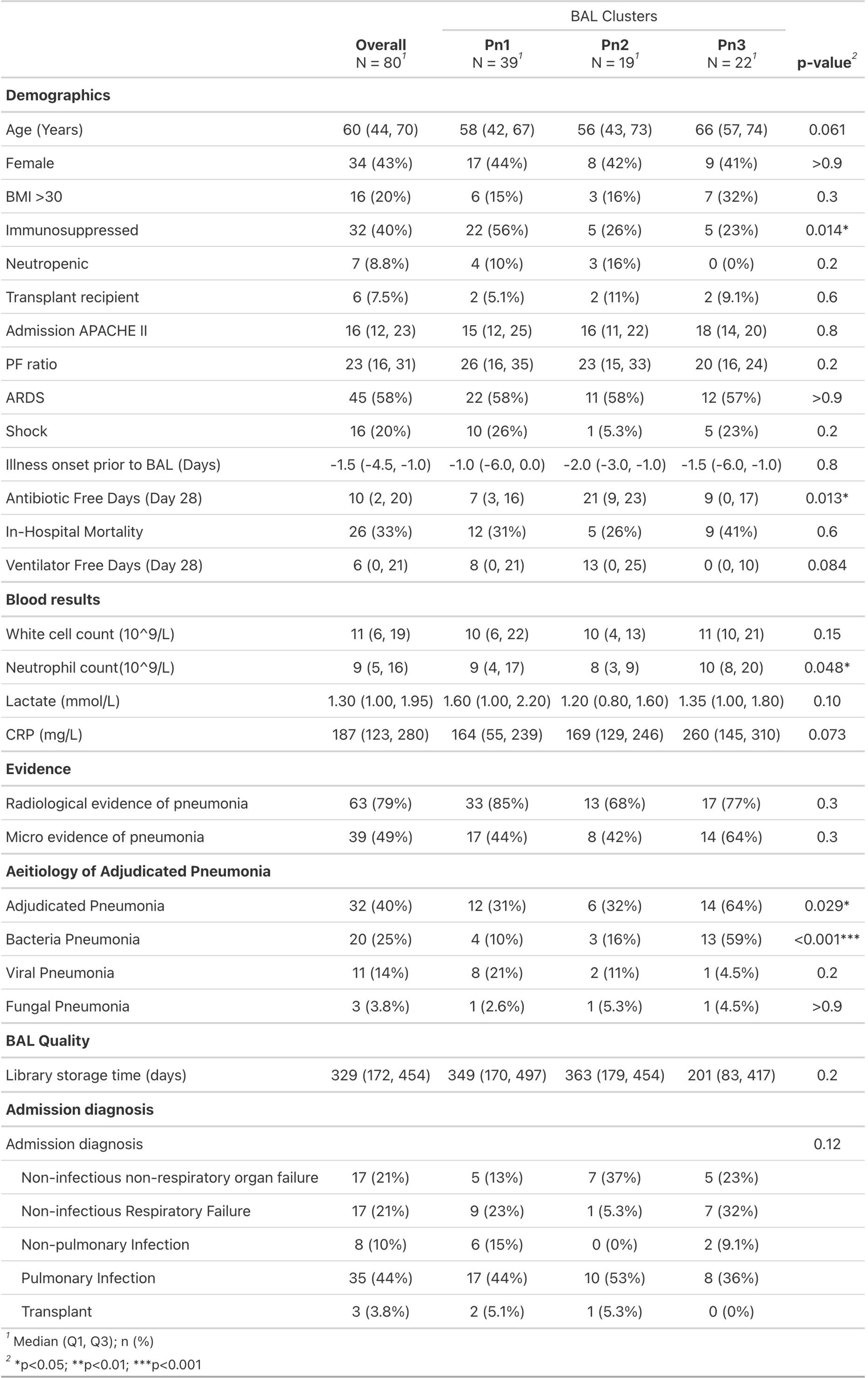
– Clinical and demographic features of the overall cohort and individual Pneumotypes. Continuous values shown as median and interquartile range, categorical by n (%)

To identify distinct pulmonary sub-phenotypes, we clustered patients based on their alveolar gene expression. Following sequencing of RNA from bronchoalveolar lavage and variance-stabilisation transformation[20] the 10% most highly variable genes were clustered using the agglomerative hybrid hierarchical k-means algorithm[21]. This identified three clusters of patients with distinct pulmonary endotypes (Figure 1A, Extended data Figure 1A-C), termed Pneumotypes 1, 2 and 3 (Pn1, 2, 3). Notably, all Pneumotypes displayed similar severity of respiratory failure, with the proportion with ARDS consistent (58%) across all three. Pn1 was enriched for immunosuppression (Figure 1B, Table 1), whilst Pn3 was enriched for bacterial pneumonia (Figure 1C, Table 1). Notably, neither of these features were exclusive to any Pneumotype, with Immunosuppression found in 26% and 23% of Pn2 and 3 respectively. Bacterial pneumonia was found in 10% and 16% of Pn1 and 2 respectively. Onset location in community or hospital were evenly distributed across the three (Figure 1D).

**Figure 1:**
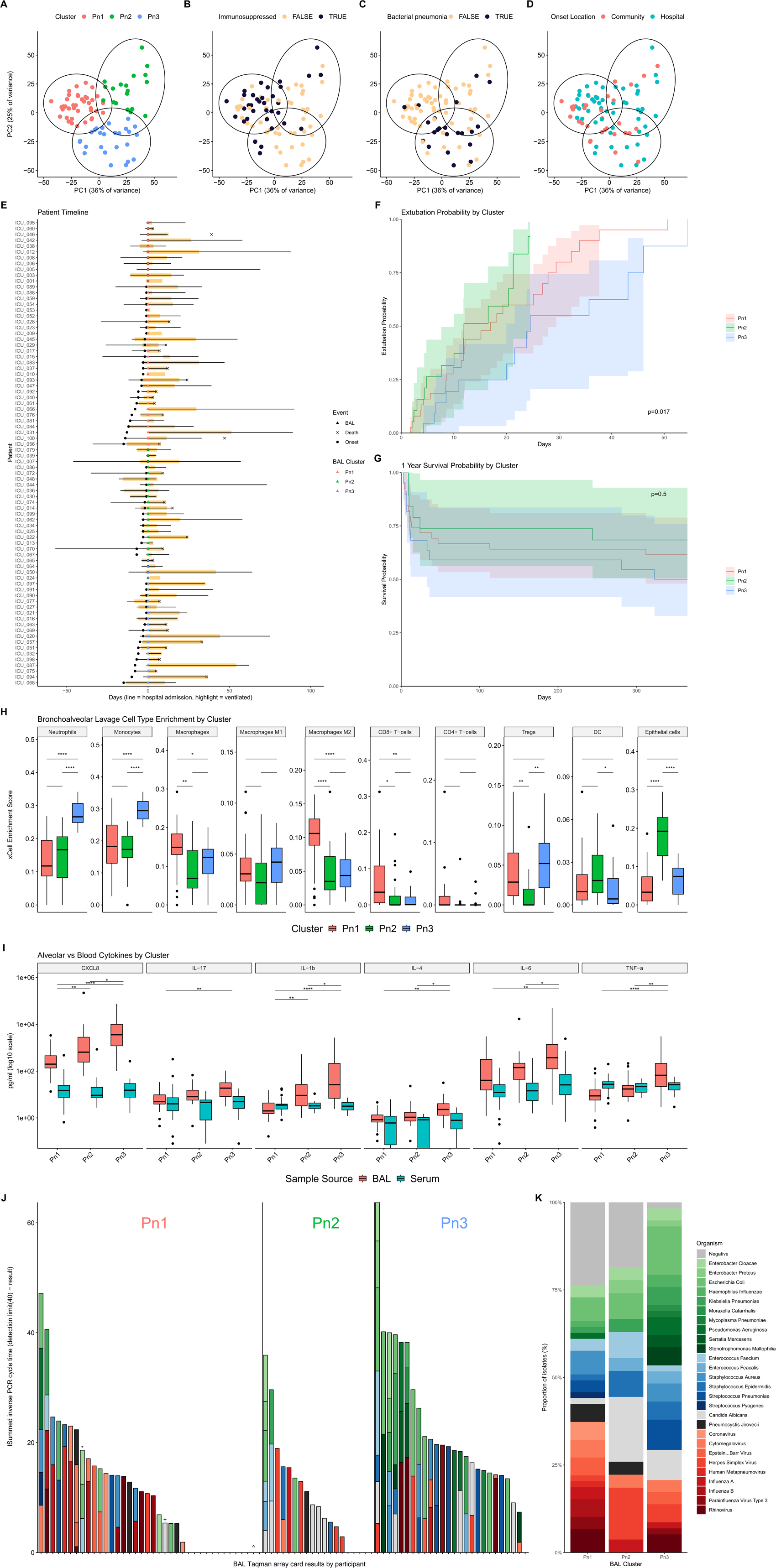
Clinical, inflammatory protein and microbiological features of Pneumotypes. First two principal components of the highly variable genes from bronchoalveolar lavage plotted with individuals coloured by **A** Pneumotype cluster, **B** immunosuppressed state, **C** adjudicated bacterial pneumonia and **D** onset location. **E** Patient timelines sorted by Pneumotype and time from illness onset to lavage. Triangle, coloured by Pneumotype, indicates lavage (red Pn1, green Pn2, blue Pn3) Solid line shows hospital admission, yellow highlight indicates the period of ventilation. Hospital admission line omitted from patients with pre-lavage hospital stays longer than 150 days. **F** Kaplan-Meier curves for time to extubation, censoring for death prior to extubation, shaded areas indicate 95% confidence intervals p-value by log-rank test. **G** Kaplan-Meier Curves for Survival to 1 year, shaded areas indicate 95% confidence intervals, p-value by log-rank test **H** Estimated cellular composition from bulk RNA deconvolution by xCell. **I** Comparison of lavage and plasma inflammatory protein concentrations by Pneumotype. **J** Pathogen TaqMan array card (TAC) results per patient, *denotes Citrobacter on sequencing but not culture or TAC, +denotes Rhinovirus on clinical PCR test but not TAC, ^ denotes Staph. Epidermidis >10^4^ CFU on culture and sequencing with negative TAC. **K** Pathogen TaqMan array card (TAC) detections summarised by Pneumotype. * adjusted p<0.05, **<0.01, ***<0.001

Pneumotypes did not appear to reflect when the patient was sampled relative to disease onset (p=0.8, Figure 1E, Extended data figure 2A), suggesting that these Pneumotypes were not different phases of disease evolution. In four cases patients were sampled twice, three during the same episode of pneumonia, demonstrating small positive migrations in principal component 2 (PC2) away from Pn3 (Extended Data Figure 2B), whilst the fourth patient was sampled during two different ICU admissions with two distinct Pneumotypes. Pn2 demonstrated significantly faster resolution of respiratory failure with shorter time to extubation when compared to Pn3 (HR for successful extubation relative to Pn2 [95% CI, P]: Pn1=0.64 [0.32-1.25, p=0.2], Pn3=0.31 [0.14-0.71, p=0.006], Fig 1F). Although the point estimate for 1-year mortality was lower in Pn2 controlling for age, the differences did not achieve statistical significance (HR for 1-year mortality relative to Pn2 [95% CI, P]: Pn1=1.34 [0.52-3.47, p=0.5], Pn3=1.44 [0.52-3.94, p=0.5], Age 1.03 [1-1.05, p=0.036], Figure 1G). The more extensive use of antimicrobials (Table 1) in Pn1 and 3 also suggest ongoing pulmonary and systemic inflammation in these Pneumotypes.

**Figure 2.**
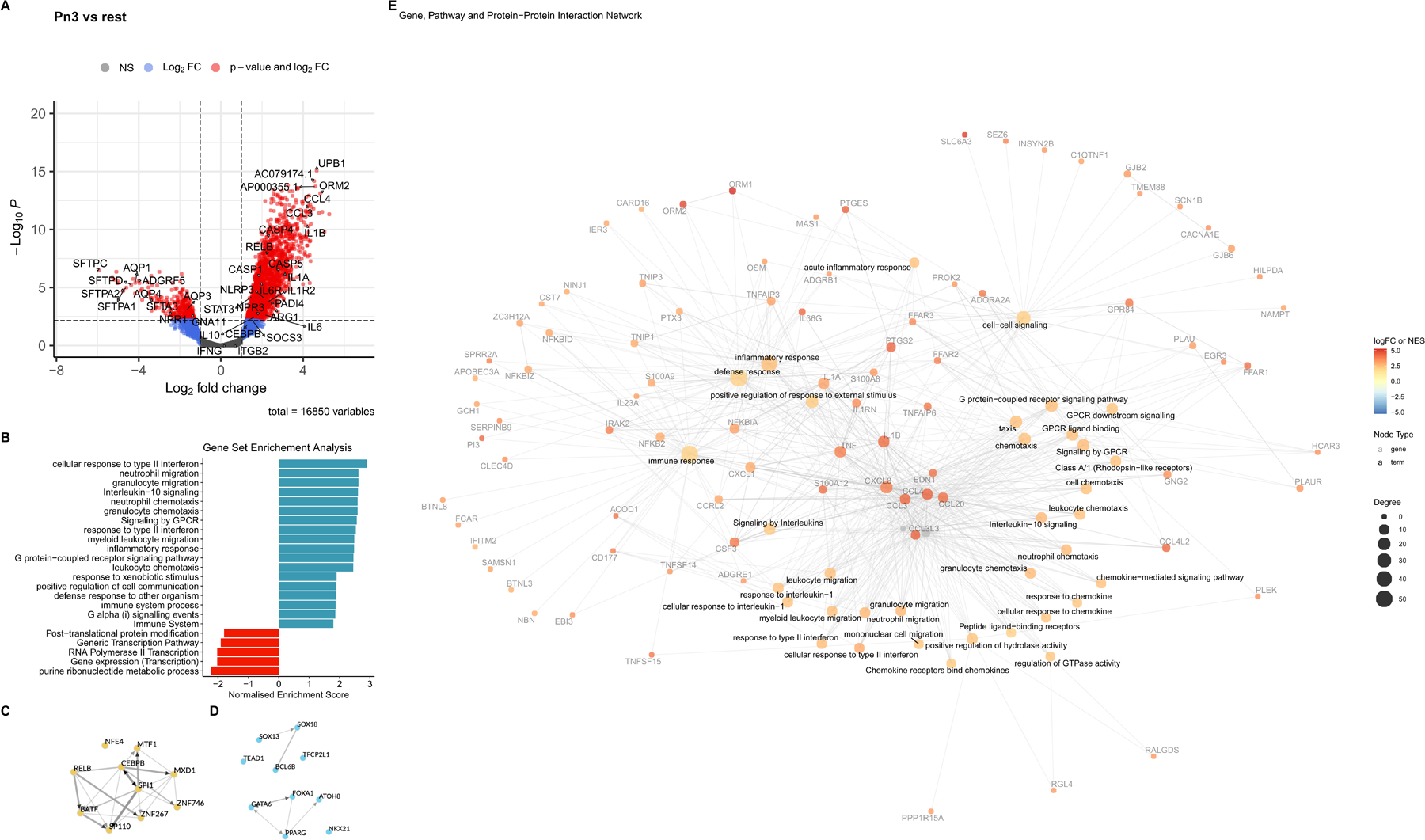
Gene transcript, pathway and transcription factor enrichment in Pneumotype 3. **A** Volcano plot of differentially expressed genes between Pn3 and the other two Pneumotypes. Each point represents a gene, those coloured in red are below adjusted p-value 0.05 **B** Gene Set Enrichment Analysis (GSEA) of Gene Ontology Biological Pathways and ReactomePA of genes differentially expressed between Pn3 and Pn1 and 2. **C** Transcription factor enrichment for differentially expressed genes in Pn3 compared to Pn1 and 2 with increased abundance and **D** decreased abundance genes derived from Chea3. **E** Pn3 Graph Network of overlapping genes from mapped pathways meshed with high confidence protein-protein interactions from StringDB (confidence score >80%). Colour denotes differential expression log-fold change for genes, or network enrichment score for pathways, and size the degree (number of network connections) for each node. Pn3 network was extremely densely overlapping relative to Pn1 and Pn2 and was therefore filtered to show just the 30 most significant mapped pathways for interpretability.

We examined the differential cellular make-up of each Pneumotype by xCell bulk RNA deconvolution[22] (Figure 1H) and conventional cytology (Extended Data Figure 3A-I). Pn1 was characterised by an expanded macrophage population, which was determined by an increased representation of macrophages labelled ‘M2’, alongside expanded regulatory CD4+ T-cells (T_reg_) and cytotoxic CD8 T-cells. Pn3 demonstrated increased infiltrating peripheral blood immune cells (neutrophils and monocytes) and T_reg_. Pn2 showed expanded epithelial and dendritic cells with an intermediate representation of macrophages and neutrophils. Conventional cytology identified a comparable pattern of neutrophils and macrophages across Pn1-3 (Extended Data Figure 3C,E), although epithelial cells were seldom identified.

We measured the concentrations of 48 inflammatory proteins, with 32 showing significantly elevated concentrations in the lavage of Pn3, relative to Pn1(Figure 1I). Pn2 had intermediate inflammatory protein levels except for CXCL1(GROα) which was highest in Pn2, demonstrating a generally U-shaped relationship between lavage inflammatory protein levels and duration of ventilation. Notably, the concentrations of plasma inflammatory proteins were comparable across all three Pneumotypes, with only IL-1ra demonstrating a significant difference. Plasma and BAL protein concentrations were mostly weakly or very weakly autocorrelated. The strongest autocorrelation was observed for IL-6 (r=0.59, Padj=1e-7), with MCP-3 (r=0.52), TNF-b and G-CSF (r=0.51), IP-10 (r=0.50) and CXCL1 (r=0.45) also showed moderate autocorrelation.

Each sample was assessed for nucleic acids associated with respiratory pathogens on a 52-organism TaqMan Array Card[18] (Figure 1J-K). These data were comparable with pathogen data extracted from metagenomic sequencing[18]. Pn3 had the highest proportion of pathogenic and non-pathogenic bacteria, with Pn1 being comprised more commonly of viral infections and samples with no pathogens detected. Pn2 had an increased proportion of low-pathogenicity organisms (*Candida* spp., *Enterococci* and coagulase-negative *Staphylococci*); however, once again no single organism type was exclusive to a given Pneumotype, and all Pneumotypes could be found in patients without an identified respiratory pathogen.

### Identifying mechanistic drivers of Pneumotypes Pneumotype 3 is characterised by inflammasome activation, expansion of immature neutrophils and impaired alveolar fluid clearance

To understand the mechanisms that underpin the Pneumotypes we examined differentially expressed genes (DEGs) between Pneumotypes in a One-vs-Rest manner. Following adjustment for age, sex, library storage time and lavage return volume, 2,411 genes were differentially expressed in Pn3 vs Pn1 and 2 (Figure 2A). Gene set enrichment analysis (GSEA) identified innate immune responses, neutrophil chemotaxis and type II interferon responses as among the most highly enriched pathways (Figure 2B). Transcription factor (TF) enrichment for up- and down-regulated gene expression using Chea3[23] (Figure 2C, D) identifies a network of proinflammatory transcription factors, including RELB and NFKB2, with enrichment of downstream genes including NLRP3, Caspases 1,4,5, IL-1β and IL-6 (Figure 2A and E) consistent with inflammasome activation and the high concentrations of alveolar cytokines identified (Figure 1I). Alveolar neutrophilia was found in both deconvolution and cytology (Figure 1H, Extended Data Figure 3E), with the enrichment of genes IL1R2, PADI4 and transcription factor CEBPB implying an expansion of immature neutrophils with reduced antimicrobial function[24]. Key mediators of emergency granulopoiesis, IL-6 and G-CSF[25] were enriched at transcript and protein level, with both correlating with plasma levels, providing a link between the lung and the bone marrow release of immature neutrophils. Monocytes are key for sustained neutrophil recruitment[26] and monocytes are also expanded in Pn3 (Figure 1H) as are the monokines CCL3 and CCL4 at both transcript and protein level (Figure 2A) Although Pn3 is characterised by immune activation and infiltration of peripheral blood leucocytes there was also evidence of concurrent immunoparesis. The elevation of counterregulatory cytokines IL-10 and IL-1RA (Figure 2A), expanded T_reg_ (Figure 1H) and enhanced expression of conventional T-cell inhibitor Arginase-1, neutrophil inhibitory C5a-receptors (C5aR1 and 2) and negative co-stimulatory molecule CD274 (PDL1) (Figure 2A) are all indicative of simultaneous activation of counterregulatory pathways.

Inspection of the downregulated transcripts in Pn3 (Figure 2A) identified genes involved in fluid clearance and alveolar surfactant function. Aquaporins (AQP) 1,3 and 4 were downregulated alongside atrial natriuretic peptide receptor 1 (NPR1), all of which play important roles in fluid clearance following acute lung injury[27, 28]. Also notable amongst the suppressed transcripts were the surfactant proteins (SFTPC, B, D, A1, A2 and SFTA3) alongside upstream receptor ADGRF5[29] and intermediate signaller GNA11[30]. Loss of surfactant proteins are an established feature of acute lung injury[31].

### Pneumotype 1 demonstrates macrophage polarization, epithelial cytopathy and T-cell mediated pathology

Differential expression for Pn1 identified 1,868 DEGs which were disproportionately down-regulated (Figure 3A). GSEA reveals activation of stress response pathways with p53 signal transduction and MAPK signalling alongside lipid metabolism changes. Downregulated pathways were predominantly those involved in epithelial function and ciliated cell activity (Figure 3B). The Chea3 analysis of upregulated transcription factors in Pn1 indicated several lipid metabolism pathways involved in alternatively activated macrophage function, with enrichment of PPARγ NR1H3 and NR1H4[32, 33] (Figure 3C). Alongside these transcription factors were multiple genes associated with alternatively activated tissue resident macrophages including CD36, MCR1, CCL18, FABP4, FBP1, MSR1 and RBP4[34] (Figure 3A). These features are all consistent with the xCell deconvolution demonstrating expansion of ‘M2 labelled’ macrophages (Figure 1H). Transcription factors associated with down-regulated genes include FOXJ1 and ELF3 (Figure 3D), both of which are involved in epithelial repair and indicate suppression of these pathways[35, 36].

**Figure 3.**
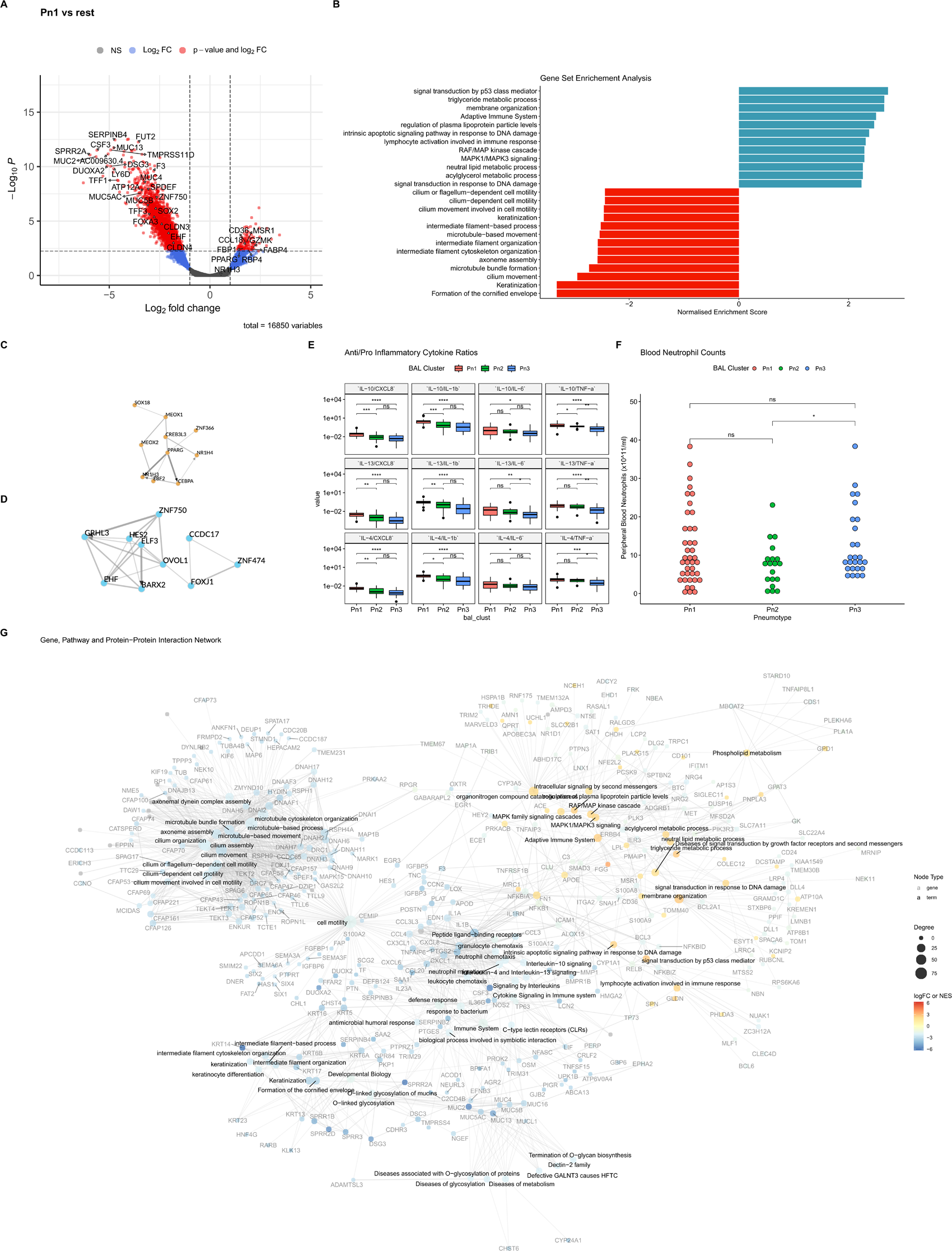
Gene transcript, pathway and transcription factor enrichment in Pneumotype 1. **A** Volcano plot of differentially expressed genes between Pn1 and the other two Pneumotypes. **B** Gene Set Enrichment Analysis of Gene Ontology Biological Pathways and ReactomePA. **C** Transcription factor enrichment for increased and **D** decreased abundance genes derived from Chea3. **E** Anti-inflammatory to pro-inflammatory lavage cytokine ratios across the Pneumotypes, indicating a relative increase in IL-4 and IL-10 in Pn1 **F** Peripheral blood neutrophil counts by Pneumotypes. **G** Pn1 Graph Network of overlapping genes from mapped pathways meshed with high confidence protein-protein interactions from StringDB (confidence score >80%). Colour denotes differential expression log-fold change for genes, or network enrichment score for pathways, and size the degree (number of network connections) for each node. * adjusted p<0.05, **<0.01, ***<0.001.

As cells are thought to respond to relative levels and changes in cytokine concentrations, rather than absolute levels[37, 38], we examined relative levels of pro and anti-inflammatory/alternative activation polarising cytokines, finding IL-10, 13 and 4 were elevated relative to CXCL8, IL-1β and TNF-α but not IL-6 (Figure 3E).

Pn1 demonstrated low levels of bronchoalveolar neutrophils (Figure 1I, Extended Data Figure 3E), however peripheral blood neutrophil counts varied considerably and this Pneumotype could develop in settings of both frank neutropenia and marked blood neutrophilia (Figure 3F). Although the expansion in CD8+ T-cells was noted in deconvolution (Figure 1H), neither the TF mapping nor DEGs identified a clear T-cell signature. However, Granzyme K was enriched in Pn1 (Figure 3A).

Examining the specific genes downregulated in Pn1 and their associated TFs points to potential mechanisms of epithelial injury and consequent loss of barrier and gas exchange functions. This epithelial barrier damage is evidenced by the increased proportion of red blood cells in the lavage of these patients (Extended Data Figure 3G) and high rates of severe respiratory failure (Table 1). The suppression of mucus production and processing genes (ATP12A, FUT2, MUC4, MUC5AC, MUC5B) alongside mucus-related transcription factors (SPDEF, FOXA3 and SOX2) and mucus components (TFF1 and TFF3) indicate a loss of this important barrier component[39]. The loss of tight junction components claudin 3 and 4 (CLDN3-4) (Figure 3A,G) also points to impaired barrier function. Tissue factor (F3), the absence of which results in alveolar haemorrhage[40], was also suppressed alongside the epithelial repair transcription factors ZNF750 and EHF (Figure 3A).

### Epithelial repair and endothelial barrier function with a balanced inflammatory response characterise Pneumotype 2

Pn2 was associated with the best outcomes and the fastest time to extubation, a signal towards possibly lower mortality and the least use of antimicrobials in the time following investigation (Figure 1F-G, Table 1). Although deconvolution indicates an expansion in epithelial cells (Figure 1H), and DEG analysis points to genes involved in cilial assembly and function (Figure 4A,B), the number of epithelial cells detected by cytology (Extended Data Fig 3A) was minimal. Differential expression and GSEA (Figure 4 A,B) indicate cell motility and cilial processes. TF mapping by Chea3 indicates enrichment of FOXA1, FOXJ1, EHF, and ELF3, which all have established roles in respiratory epithelial repair [35, 36] as well as IRF-6, an epithelial restricted interferon response protein with both barrier integrity and immune response functions [41] (Figure 4C). The TFs associated with down-regulated genes include CEPBE and SPI1 (Figure 4D), which point to the presence of mature neutrophils in contrast to the immature neutrophils seen in Pn3.

**Figure 4:**
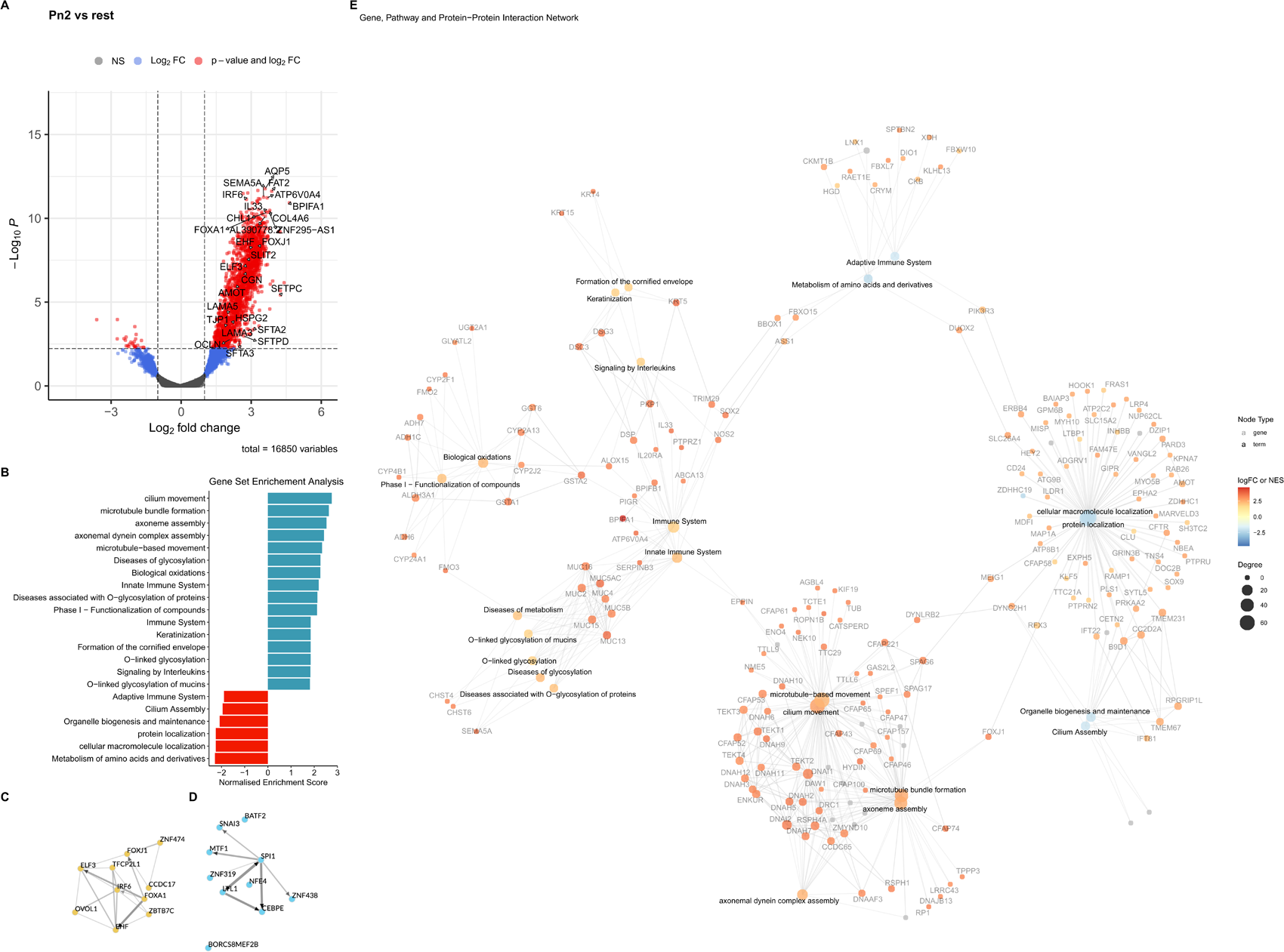
Gene transcript, pathway and transcription factor enrichment in pneumotype 2. **A** Volcano plot of differentially expressed genes between Pn2 and the other two Pneumotypes. **B** Gene Set Enrichment Analysis of Gene Ontology Biological Pathways and ReactomePA. **C-D** Transcription factor enrichment for increased abundance genes (**C**) and decreased abundance (**D**) differentially expressed in Pn2, derived from Chea3. **E** Pn1 Graph Network of overlapping genes from mapped pathways meshed with high confidence protein-protein interactions from StringDB (confidence score >80%). Colour denotes differential expression log-fold change for genes, or network enrichment score for pathways, and size the degree (number of network connections) for each node

Manual review of gene expression also identified upregulated endothelial gene expression in this Pneumotype (Figure 4A). SLIT2 and ROBO, which play a key role in preserving endothelial barrier function in infectious pulmonary insults[42] were upregulated alongside tight junction proteins TJP1, OCLN, CGN and tight junction regulators AMOT and JUP1 and basement membrane components HSPG2, LAMA3 and LAMA5 (Figure 4A).

The gene-pathway-protein-protein interaction network (Figure 4E) also indicated innate immune activity, which is consistent with the moderate levels of infiltrating immune cells identified by deconvolution and cytology as well as the intermediate levels of lavage cytokines and inflammatory proteins. Pn2, therefore appears to be the most adapted Pneumotype of the three identified, with a balanced immune response and pro-resolution epithelial and endothelial responses.

### Compartmentalisation of lung responses

We next investigated gene expression in peripheral blood to determine whether Pneumotypes were associated with systemic responses. After filtering, 76.8% of expressed genes (13,540 common genes out of 17,625 total, 2,586 unique to blood and 1,499 unique to BAL) were identified in both blood and bronchoalveolar lavage. Differential gene expression in blood did not identify any significantly differentially expressed genes in either Pn1 or Pn2, and only 100 genes in Pn3 which mapped to neutrophil degranulation [p=1.043e-15] and innate immune system [p=6.346e-8] Reactome terms. A weighted gene correlation network analysis (WGCNA) (Figure 5A upper three rows) demonstrated similarly bland responses when segregated by Pneumotype. Further, no plasma inflammatory proteins differed significantly between Pneumotypes compared to 35/48 proteins measured in lavage (Figure 1I). Thus, identification of Pneumotypes from blood is unlikely to be feasible. For completeness we created a WGCN for the clusters derived from BAL (Extended Data Figure 4A). This identified 9 modules with marked differences between the pneumotypes with pathway mapping results consistent with the DEG analysis above (Extended Data Figure 4B-J).

**Figure 5:**
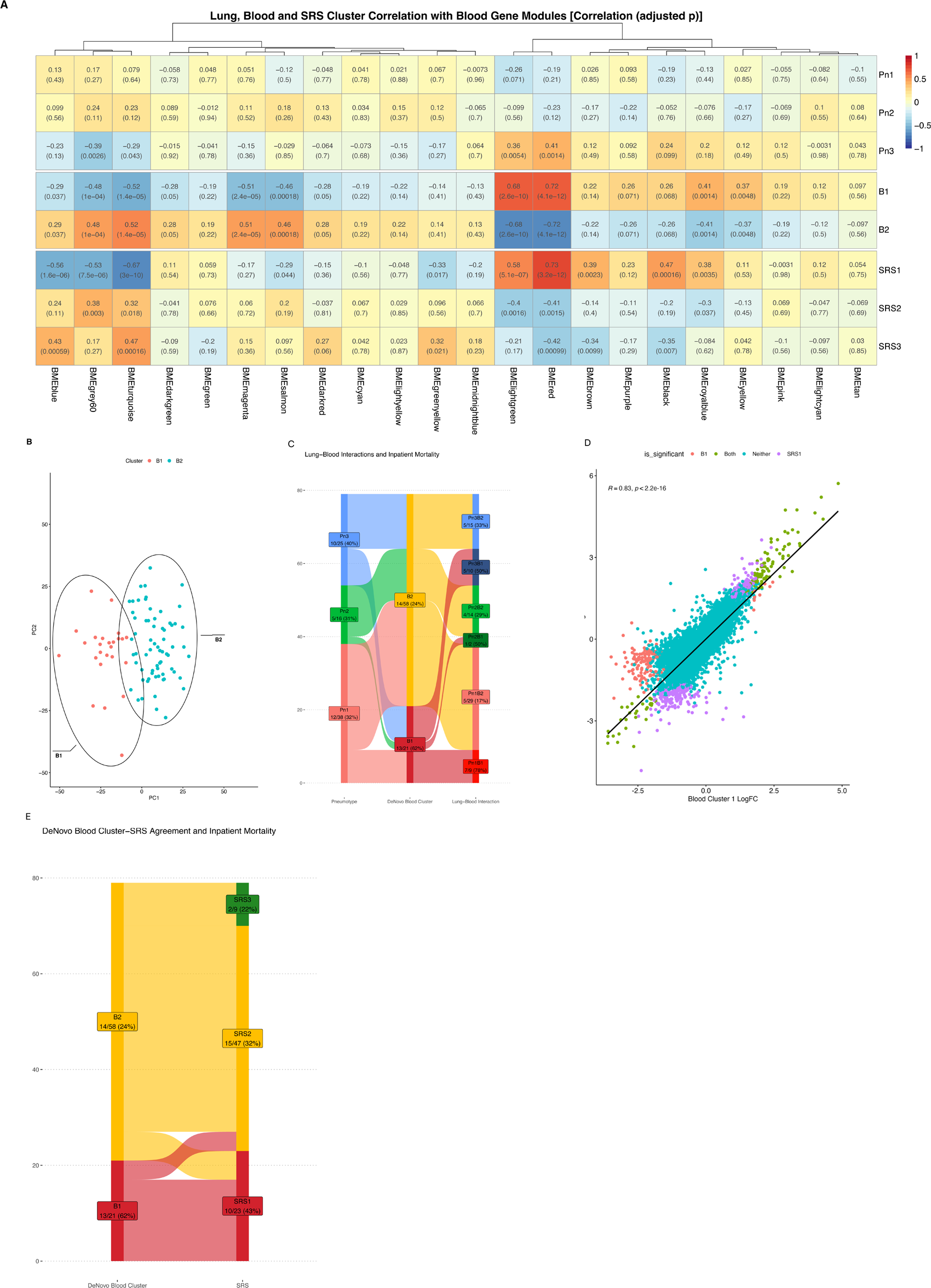
Transcriptional features of the blood and Pneumotype-blood interactions. **A** Heatmap of lung and blood cluster correlations with blood gene co-expression modules. Upper 3 rows show Pneumotypes(Pn1-3), middle two Blood clusters (B1 and 2) and bottom SRS groups (SRS1-3). **B** First two principal components of the highly variable genes from blood plotted with individuals coloured by blood cluster. **C** Alluvial plot of lung and blood cluster interactions and in-hospital mortality [n/total (%)]. **D** Correlation in Log_2_-fold change of all detected genes in blood between participants assigned to Blood 1 and SRS1 phenotypes. Colours indicate FDR <0.05, red Blood1 only, green both Blood1 and SRS1, purple SRS1 only and turquoise neither. **E** Alluvial plot of Blood and SRS assignments, percentage indicates inpatient mortality.

De-novo clustering of the blood transcripts by HKMC identified two clusters (Figure 5A middle two rows, 5B,Extended Data Figure 5A-C) with divergent outcomes and clinical features (Extended Data Table 1). The two clusters, termed Blood 1 (B1) and Blood 2 (B2), had distinct enrichment patterns with Blood 1 showing highly upregulated coagulation (greenyellow) and innate response, neutrophil degranulation and TLR activation(pink) and moderately upregulated interferon response (tan) blood co-expression modules. Downregulated modules included RNA metabolic processes (turquoise), mitochondrial translation (lightgreen), nitrogen compound metabolic processes(blue) and erythrocyte homeostasis (magenta). (Figure 5A).

The distribution of blood phenotypes was uneven across the Pneumotypes, with Pn3 having proportionately more of the maladaptive Blood 1 phenotype, although 40% co-occurred with Blood 2. Dyads formed by different Pneumotypes and blood phenotypes reveal divergent mortality outcomes (Figure 5C), with the most notable difference seen in Pn1.

Our Blood clusters were reminiscent of the previously described Sepsis Response Syndrome Signatures (SRS)[15]. Examination of differentially expressed genes between Blood 1 and 2 and SRS 1 and 2 identified a high degree of correlation (r=0.83, p<0.001) (Figure 5D) reflected in similar patterns in WGNA module correlation (5A, bottom 5 rows) and significant correspondence between allocations (B1-SRS1 and B2-SRS2 (5E)). Although both our Blood types and SRS assignments identified greater severity of illness in Blood 1/SRS1 this was more pronounced in Blood clusters and was reflected in a significant difference in mortality (5E, Extended Data Table 1).

### Validation of Pneumotypes

Although publicly available datasets that are directly comparable could not be identified, two cohorts of adult patients with clinically suspected pneumonia were available with sufficiently similar data to allow comparison. Langellier and colleagues collected tracheal aspirate (TA) from patients with suspected pneumonia and undertook bulk host RNA transcriptomics[19]. Grant and colleagues collected bronchoalveolar lavage from patients with microbiologically confirmed pneumonia and controls and undertook flow-cytometric cell identification alongside bulk RNA sequencing of sorted alveolar macrophages[43].

Examination of the endotracheal aspirates revealed an unsurprising paucity of alveolar cells, specifically lacking signals for macrophages and lymphocytes (Figure 6A,B). Clustering metrics indicate two clusters (Extended Data Figure 5C-E), rather than the three present in bronchoalveolar samples. To compare the similarity of BAL and TA clusters, we generated a consensus gene co-expression network between Langelier’s and our cohort. Using this approach we identified two phenotypes with similar pathways enriched to those seen in Pn2 and Pn3 in our cohort (Figure 6C), termed TA1 and TA2. TA2 shared clinical and transcriptomic features with Pn3 (Figure 6A, 6C, Extended Data Table 2), and was enriched for bacterial pneumonia. The top group of modules in Figure 6C (purple to yellow) map to innate immune responses and showed high correlation between Pn3 and TA2 with corresponding downregulation of the metabolic modules (black to pink). Pn2 and TA1 were clustered together with high correlation of cilial assembly modules (brown), but TA1 also shared features with Pn1 with respect to upregulation of metabolic modules (Black-Pink). xCell-derived cellularity indicated increased neutrophil, monocyte and T_reg_ numbers in TA2 (Figure 6A). TA1 most closely resembles Pn2 with an enriched epithelial repair signal module, T_reg_ restriction and reduced innate response modules and neutrophillia relative to TA2/Pn3. However, the inability to identify Pn1 through the paucity of macrophages (Figure 6B) indicates a significant limitation in tracheal aspirate transcriptomics for the assessment of the pulmonary host response.

**Figure 6:**
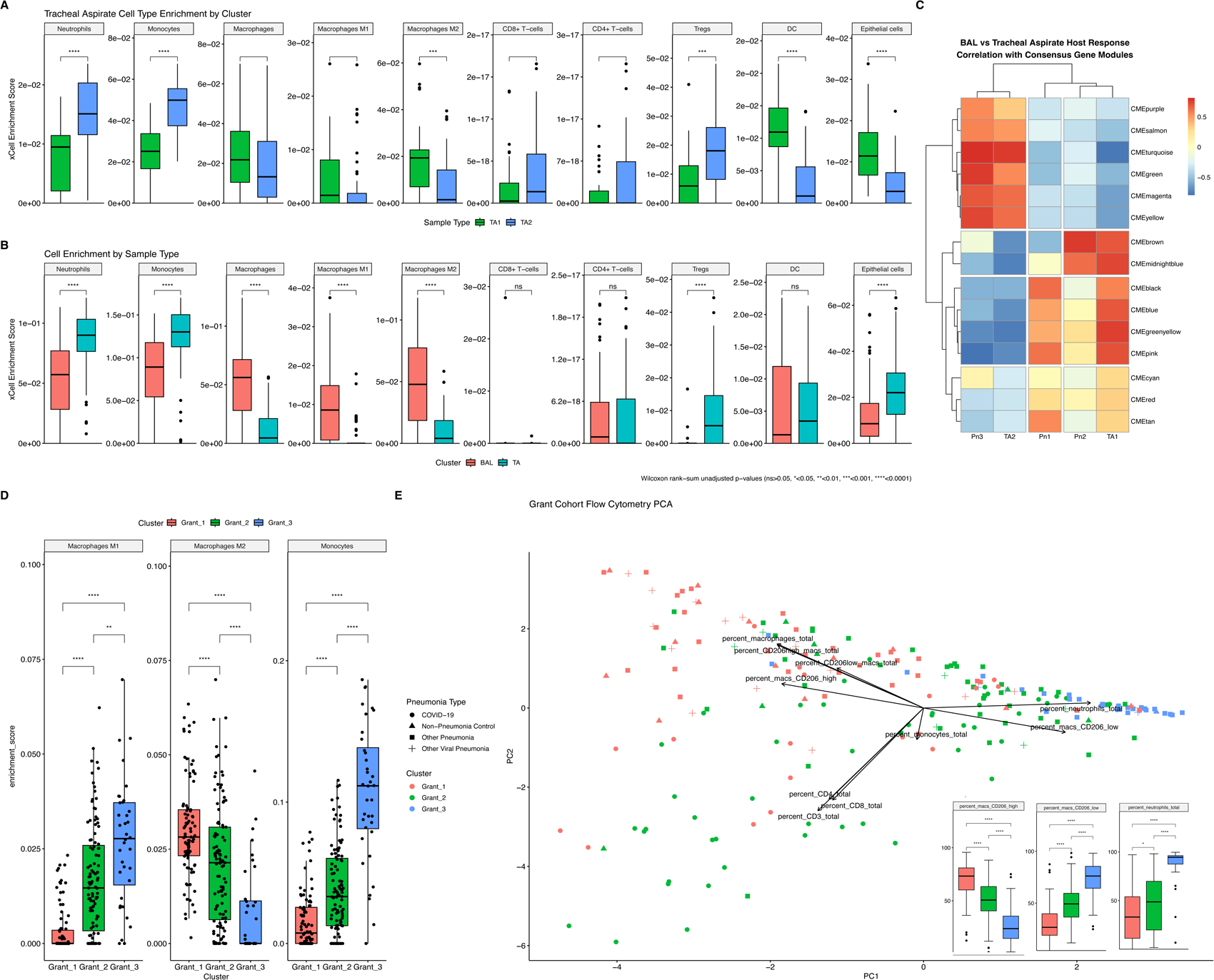
Validation of Pneumotypes in External Datasets. **A** Estimated cellular proportions by xCell bulk RNA deconvolution of tracheal aspirate (TA) clusters TA1 (green) and TA2 (blue) from Langelier et al [17]. **B** Comparative estimated cellular proportions of TA from Langelier et al and BAL from bronchoalveolar lavage dataset reported in this paper, by xCell RNA deconvolution. **C** Tracheal aspirate from Langelier dataset and our bronchoalveolar cell consensus gene co-expression modules derived from weighted gene co-expression network analysis (WGCNA) hierarchically clustered by similarity. **D** Differential cellularity of BAL estimated by xCell bulk RNA deconvolution from flow-sorted Macrophages from Grant cohort[43]. **E** Principal Component Analysis of flow cytometry proportions in Grant et. al. cohort labelled for cluster (Grant 1 red, Grant 2 Green, Grant 3 Blue) and diagnosis (circle COVID-19, triangle non-pneumonia control, square -bacterial pneumonia, cross non-COVID viral pneumonia). Inset shows summary of flow cytometric identification of CD206+ Macrophages, CD206-Macrophages and neutrophils.

By contrast, the Grant dataset lacked the ability to detect Pn2 through lack of total bronchoalveolar cell bulk RNA sequencing and no epithelial cells identified by flow cytometry. Clustering the sorted macrophage RNA sequencing identified three clusters (Extended data Figure 5F-H), with xCell deconvolution indicating M1/M2 macrophage polarisation and monocyte enrichment consistent with Pn1 and Pn3 (Figure 6D). Examining the flow cytometric counts with each cluster confirmed the macrophage polarisation (Figure 6E) and identified enrichment for neutrophils in the ‘M1’ biased cluster and paucity of neutrophils in the ‘M2’ biased cluster (Inset box and whisker plots, Figure 6E). 27 out of 36 (75%) of Grant cluster 3 had bacterial pneumonia, consistent with Pn3 (Extended data Table 3). The third cluster (Grant 2) in the Grant data set was enriched for T-lymphocytes and has disproportionate numbers of patients with COVID-19. This is likely to be the COVID-specific phenotype Grant and colleagues identified in their original report[43].

## Discussion

Using bulk RNA sequencing on bronchoalveolar fluid we have identified three phenotypes in the lungs of patients with lung injury and suspected pneumonia. These phenotypes were reflected in the differential immune cell populations and inflammatory proteins. These phenotypes are compartmentalised to the lungs, are non-synonymous but interact with the peripheral blood immune phenotype and can be identified in external datasets drawn from pulmonary samples. Each of these Pneumotypes is underpinned by distinct mechanisms and implies differential responses to therapies. We have been able to validate each of the Pneumotypes in other data sets [17,37]. They also bear comparison to recently described sub-phenotypes in the lungs of children with lung injury following bone-marrow transplantation[44]. Zinter and colleagues identified four sub-phenotypes, with differential alveolar cell types. One of these featured high levels of bacteria and neutrophils, like Pn3, although the remaining 3 sub-phenotypes did not match with those identified in our study. The difference in age (children vs adults) and being bone marrow transplantation recipients may explain these divergent findings.

Each of the Pneumotypes contained both patients with and without confirmed pneumonia, implying common mechanisms underpinning lung injury arising from different mechanisms. This observation provides insight into previous failures to identify specific markers that distinguish pneumonia from other forms of lung injury[4]. The non-synonymous nature of the blood and broncho-alveolar phenotypes sounds a note of caution regarding the use of blood phenotypes alone to guide therapy[17].

Pneumotype 3 is perhaps the most immediately recognisable Pneumotype, with its neutrophil dominant cytology, impairment of alveolar fluid clearance and loss of surfactant it is closest to the classical description of pneumonia and ARDS pathophysiology[31]. In findings reminiscent of Kwok et al’s description of the neutrophil phenotype in SRS1[24], we found signals for enrichment of immature neutrophils in this setting. Immature neutrophils are known to have impaired antimicrobial functions but enhanced degranulation and consequent tissue toxicity[25, 45]. The non-synonymous relationship between Pn3 and Blood 1 (the latter being similar to SRS1) suggests that recruitment of immature neutrophils to the lungs in Pn3 may be selective and specific rather than simply reflecting peripheral blood left-shifted granulocytosis.

The lung monocyte-neutrophil co-recruitment in Pn3, with IL-6 signalling sustaining emergency granulopoesis with skewing of haematopoetic stem cells towards granulocyte production[25, 46] may form a positive feedback loop, creating a bi-stable equilibrium[47] that sustains prolonged inflammation and organ impairment that can persist after the triggering insult is removed (schematic in Extended Data Figure 6A). The phenomenon of persisting inflammation after pathogen removal is well described, but poorly understood[48–50]. Our findings suggest that, alongside adequate pathogen control, patients with Pn3 may benefit from targeted immunomodulation such as selective IL-6 blockade. Conversely, such approaches may be harmful in Pn1 and of limited therapeutic impact in Pn2.

Although Pn1 was characterised by macrophages enriched for an apparent ‘pro-resolution’ transcriptional pattern, these patients have a similar degree and severity of lung injury. This observation illustrates the phenomenon of non-neutrophil-induced lung injury. The existence of ARDS in neutropaenic patients has long been described[51, 52] but the mechanisms that underpin this syndrome have remained obscure[52]. In Pn1 viral pathogens, granzyme K release from CD8+ T-cells[53] and further, as yet unidentified, factors may induce epithelial cytopathy. This leads to epithelial disruption and lung leak as exemplified by alveolar haemorrhage, differentiating this from neutrophil-driven damage in Pn3.

Regarding the development and maintenance of Pn1, alternatively activated macrophages can exclude neutrophils from a tissue space[54]. Conversely, neutrophils themselves can induce a pro-inflammatory macrophage phenotype[55]. This phenotype may therefore arise from both an absence of peripheral blood neutrophils or polarisation of macrophages in the lungs (schematic in Extended Data Figure 6B). Pn1 therefore, appears to be maintained by tolerogenic macrophages, that are unable to clear pathogens, with consequent recruitment of CD8 cells that either alone, or in combination with macrophages and direct pathogen effects, induce cytopathic effects in alveolar epithelium.

Although the phenotypes identified in the lungs are not well reflected in the blood, there are interactions that associate with different outcomes. Notably, the “adverse” blood phenotype is enriched in patients with Pn3, potentially explaining why previous studies appear to identify distinct lung phenotypes based on blood profiling in ARDS[16]. However, the work presented here demonstrates the need to assess both these compartments to understand the immunopathology and aid prognostication. Each compartment can be assigned a phenotypic category that combines to give an overall status (e.g. Pn1B1 or Pn2B1). Blood is a liminal fluid, connecting distinct tissue beds and allowing bi-directional interactions. Therefore, a fuller appreciation of the immunopathology in pneumonia, acute lung injury and indeed sepsis more widely is likely to require examination of other tissue compartments, most notably the bone marrow.

This study has several strengths, through its inclusion of a broad range of patients with diverse range of pathogens and sites of onset we can draw inferences about commonalities and differences between these groups. The phenotypes we have identified are robust to the clustering approach used and we can identify similar Pneumotypes in external datasets. There remain several areas of uncertainty. First, although we have validated in external datasets, further validation of these Pneumotypes in a full replication cohort using the same inclusion, sampling and analysis techniques is required. Whilst the temporal relationships between disease onset and sampling, and the few serial samples we have, do not point to the Pneumotypes being features of a common pathway sampled at different times, serial sampling will be required to confirm temporal stability and understand phenotype evolution and recovery trajectories. Although we have identified differential cellularity as part of the phenotypes, the use of bulk RNA sequencing does not allow us to determine which transcriptional programs are associated with a specific cell type. This is most prominent in the epithelial and endothelial signatures in Pn2 where the cytological evidence for presence of these cells is sparse. These signatures may be driven by a small number of hypertranscriptional stem cells involved in regeneration and repair[56]. The ability of distal airway stem cells to protect against lung injury in influenza models suggests these as a potential mediator of the protective phenotype in Pn2[57]. Although we have identified three Pneumotypes, it is likely that other Pneumotypes may exist and may be identified in larger cohorts or those with distinct triggering pathologies, such as the potentially distinct T-cell driven responses in COVID-19 reported by Grant et al[43].

In conclusion, we have identified three pulmonary-confined endotypes in patients with severe pneumonia and lung injury. These phenotypes are underpinned by distinct mechanisms and have differential outcomes. The mechanisms point to different therapeutic options, as well as extending our understanding of the biology of lung inflammation in the context of severe pneumonia.

## Data Availability

All data produced in the present study are available upon reasonable request to the authors

## Tables

**Extended data Table 1:** Clinical and demographic features of the patients when clustered by blood type or SRS type[15]. Continuous values shown as median and interquartile range, categorical by n (%)

| Group |  | Blood 1, N = 21 | Blood 2, N = 58 | p-value | SRS1, N = 23 | SRS2, N = 47 | SRS3, N = 9 | p-value |
| --- | --- | --- | --- | --- | --- | --- | --- | --- |
| Demographics and clinical features | Age (Years) | 60 (51, 76) | 60 (43, 69) | 0.7 | 61 (53, 75) | 60 (43, 70) | 43 (41, 54) | 0.029* |
|  | Female | 7 (39%) | 24 (43%) | 0.8 | 11 (52%) | 16 (36%) | 4 (44%) | 0.5 |
|  | BMI >30 | 4 (22%) | 11 (20%) | >0.9 | 4 (19%) | 9 (20%) | 2 (22%) | >0.9 |
|  | Immunosuppressed | 10 (56%) | 20 (36%) | 0.14 | 10 (48%) | 17 (39%) | 3 (33%) | 0.7 |
|  | Neutropenic | 3 (17%) | 2 (3.6%) | 0.089 | 2 (9.5%) | 2 (4.5%) | 1 (11%) | 0.5 |
|  | Transplant recipient | 2 (11%) | 3 (5.4%) | 0.6 | 3 (14%) | 0 (0%) | 2 (22%) | 0.006** |
|  | Admission APACHE II | 23 (20, 27) | 14 (11, 18) | <0.001*** | 21 (17, 27) | 15 (12, 19) | 12 (10, 15) | 0.002** |
|  | PF ratio | 20 (13, 25) | 24 (17, 33) | 0.2 | 20 (13, 26) | 24 (18, 33) | 24 (17, 31) | 0.3 |
|  | ARDS | 12 (71%) | 30 (55%) | 0.2 | 13 (62%) | 24 (57%) | 5 (56%) | >0.9 |
|  | Shock | 6 (33%) | 9 (16%) | 0.2 | 4 (19%) | 10 (23%) | 1 (11%) | >0.9 |
|  | Illness onset prior to BAL (Days) | -2.5 (-3.0, -1.0) | -1.0 (-6.0, 0.0) | 0.3 | -1.0 (-3.0, -1.0) | -1.0 (-6.0, -1.0) | -2.0 (-8.0, -1.0) | 0.5 |
|  | Antibiotic Free Days (Day 28) | 3 (0, 15) | 12 (5, 20) | 0.018* | 7 (0, 18) | 11 (5, 20) | 15 (3, 18) | 0.3 |
|  | In-Hospital Mortality | 10 (56%) | 14 (25%) | 0.016* | 8 (38%) | 14 (32%) | 2 (22%) | 0.8 |
|  | Ventilator Free Days (Day 28) | 0 (0, 22) | 8 (0, 21) | 0.4 | 6 (0, 24) | 8 (0, 21) | 1 (0, 9) | 0.6 |
| Laboratory results | White cell count (10<sup>9</sup>/L) | 16 (5, 24) | 10 (7, 16) | 0.5 | 18 (6, 24) | 10 (7, 16) | 10 (6, 11) | 0.3 |
|  | Neutrophil count(10<sup>9</sup>/L) | 15 (4, 23) | 8 (6, 13) | 0.3 | 17 (5, 23) | 8 (6, 14) | 8 (4, 9) | 0.069 |
|  | Lactate (mmol/L) | 1.70 (1.45, 2.10) | 1.20 (0.98, 1.83) | 0.005** | 1.70 (1.40, 1.90) | 1.30 (1.00, 2.20) | 1.00 (0.80, 1.30) | 0.053 |
|  | CRP (mg/L) | 300 (252, 329) | 150 (84, 231) | <0.001*** | 274 (206, 318) | 170 (134, 237) | 120 (94, 158) | 0.004** |
| Evidence | Radiological evidence of pneumonia | 17 (94%) | 40 (71%) | 0.055 | 20 (95%) | 31 (70%) | 6 (67%) | 0.037* |
|  | Micro evidence of pneumonia | 11 (61%) | 26 (46%) | 0.3 | 10 (48%) | 23 (52%) | 4 (44%) | 0.9 |
| Aeitiology of Adjudicated Pneumonia | Bacteria Pneumonia | 8 (44%) | 11 (20%) | 0.06 | 9 (43%) | 10 (23%) | 0 (0%) | 0.037* |
|  | Viral Pneumonia | 4 (22%) | 6 (11%) | 0.2 | 2 (9.5%) | 7 (16%) | 1 (11%) | 0.9 |
|  | Fungal Pneumonia | 3 (17%) | 0 (0%) | 0.013* | 3 (14%) | 0 (0%) | 0 (0%) | 0.033* |
| Blood Quality | Library storage time (days) | 376 (202, 472) | 329 (111, 431) | 0.3 | 334 (201, 476) | 313 (111, 434) | 359 (200, 402) | 0.6 |
|  | Lavage volume >=200ml | 14 (78%) | 40 (71%) | 0.8 | 15 (71%) | 33 (75%) | 6 (67%) | 0.8 |
|  | Lavage return volume (ml) | 56 (31, 96) | 60 (30, 80) | >0.9 | 50 (30, 80) | 61 (34, 100) | 60 (50, 61) | 0.3 |
| Admission diagnosis | Admission diagnosis |  |  | 0.12 |  |  |  | 0.2 |
|  | Non-infectious non-respiratory organ failure | 2 (11%) | 14 (25%) |  | 2 (9.5%) | 12 (27%) | 2 (22%) |  |
|  | Non-infectious Respiratory Failure | 2 (11%) | 14 (25%) |  | 4 (19%) | 9 (20%) | 3 (33%) |  |
|  | Non-pulmory Infection | 4 (22%) | 3 (5.4%) |  | 3 (14%) | 3 (6.8%) | 1 (11%) |  |
|  | Pulmory Infection | 9 (50%) | 23 (41%) |  | 9 (43%) | 20 (45%) | 3 (33%) |  |
|  | Transplant | 1 (5.6%) | 2 (3.6%) |  | 3 (14%) | 0 (0%) | 0 (0%) |  |

**Extended data table 2:**
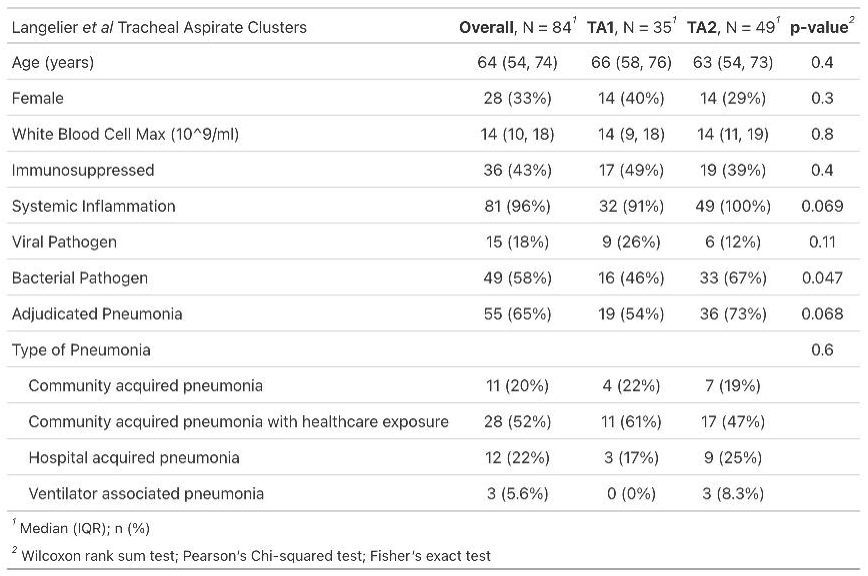
Clinical and demographic features of patients from Langellier et al[19] clustered by tracheal aspirate bulk RNA sequencing into TA1 and TA2.

**Extended data table 3:** Clinical and demographic features of patients from Grant et al[43] clustered by sorted alveolar macrophage bulk RNA sequencing into G1, G2 and G3.

| Grant et al Macrophage RNA Clusters |  |  |  |  |  |
| --- | --- | --- | --- | --- | --- |
| Characteristic | Overall, N = 167<sup>†</sup> | Grant_1, N = 59<sup>†</sup> | Grant_2, N = 78<sup>†</sup> | Grant_3, N = 30<sup>†</sup> | p-value<sup>‡</sup> |
| Age | 60 (47, 70) | 60 (43, 70) | 57 (44, 68) | 69 (60, 74) | 0.002 |
| Female | 69 (41%) | 25 (42%) | 38 (49%) | 6 (20%) | 0.025 |
| Days since intubation | 3 (1, 11) | 6 (2, 18) | 3 (1, 8) | 2 (1, 9) | 0.073 |
| Diagnosis |  |  |  |  | <0.001 |
| Bacterial Pneumonia | 79 (47%) | 22 (37%) | 33 (42%) | 24 (80%) |  |
| COVID-19 | 45 (27%) | 12 (20%) | 32 (41%) | 1 (3.3%) |  |
| Non-Pneumonia Control | 21 (13%) | 14 (24%) | 6 (7.7%) | 1 (3.3%) |  |
| Viral Pneumonia | 22 (13%) | 11 (19%) | 7 (9.0%) | 4 (13%) |  |
| Bacterial Superinfection (if Viral) |  |  |  |  | 0.007 |
| Not Viral | 102 (61%) | 37 (63%) | 39 (50%) | 26 (87%) |  |
| Primary Only | 48 (29%) | 16 (27%) | 30 (38%) | 2 (6.7%) |  |
| Superinfection | 17 (10%) | 6 (10%) | 9 (12%) | 2 (6.7%) |  |
| CD206 high Macs (% Macs) | 56 (33, 73) | 73 (61, 80) | 51 (34, 64) | 24 (14, 39) | <0.001 |
| CD206 low Macs (% Macs) | 44 (27, 67) | 27 (20, 39) | 49 (36, 66) | 76 (61, 86) | <0.001 |
| CD3+ T-cells (% total) | 5 (1, 14) | 5 (2, 13) | 7 (3, 22) | 1 (0, 2) | <0.001 |
| CD4+ T-cells (% total) | 2 (0, 6) | 2 (0, 6) | 3 (1, 9) | 0 (0, 1) | <0.001 |
| CD8+ T-cells (% total) | 2 (1, 6) | 2 (1, 5) | 4 (1, 11) | 0 (0, 1) | <0.001 |
| Neutrophils (% total) | 53 (21, 82) | 35 (14, 60) | 50 (20, 70) | 94 (85, 96) | <0.001 |
| Macrophages (% total) | 27 (7, 48) | 42 (25, 68) | 24 (10, 39) | 2 (1, 9) | <0.001 |
| Monocytes (% total) | 4.1 (2.2, 7.3) | 3.4 (1.2, 6.4) | 5.9 (3.1, 9.9) | 3.1 (2.1, 4.0) | <0.001 |
| <sup>†</sup> Median (IQR); n (%) |  |  |  |  |  |
| <sup>‡</sup> Kruskal-Wallis rank sum test; Pearson's Chi-squared test; Fisher's test with simulated p-value; Fisher's exact test |  |  |  |  |  |

## Figure legends

**Extended Data Figure 1:**
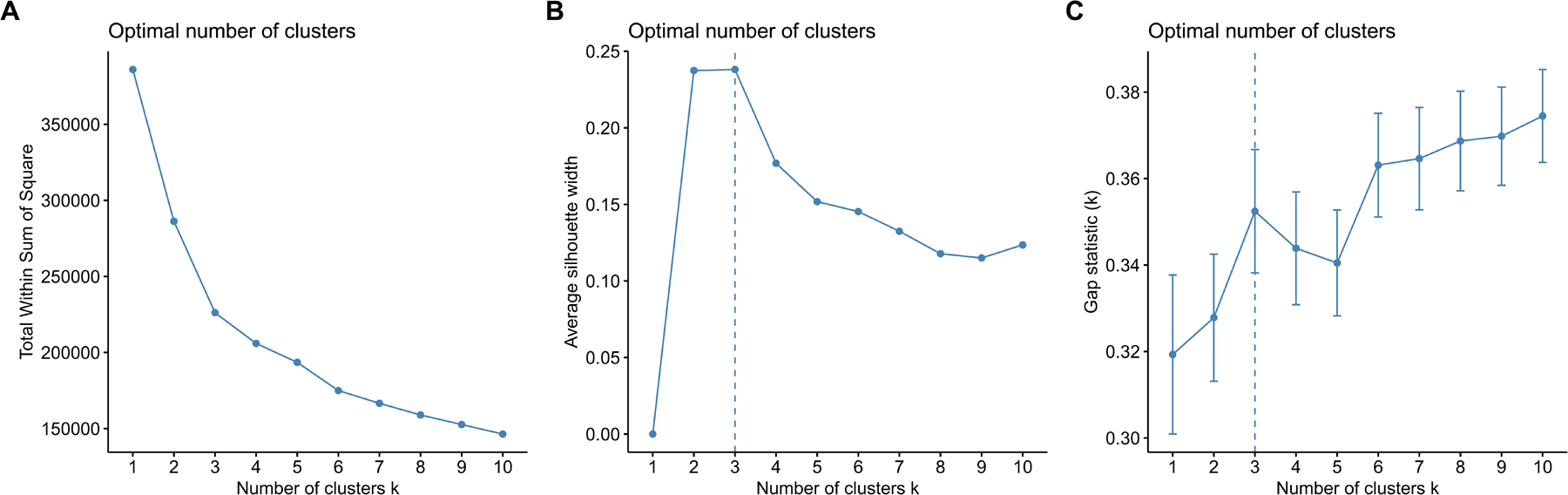
clustering metrics for BAL gene expression. **A** Elbow plot demonstrating an elbow at 3 and no further reduction in with-group sum of squares between 3 and 5 clusters. **B** Silhouette score plot with maximal scores between two and three clusters. **C** Gap statistic plot indicating a local maximum at three clusters.

**Extended Data Figure 2:**
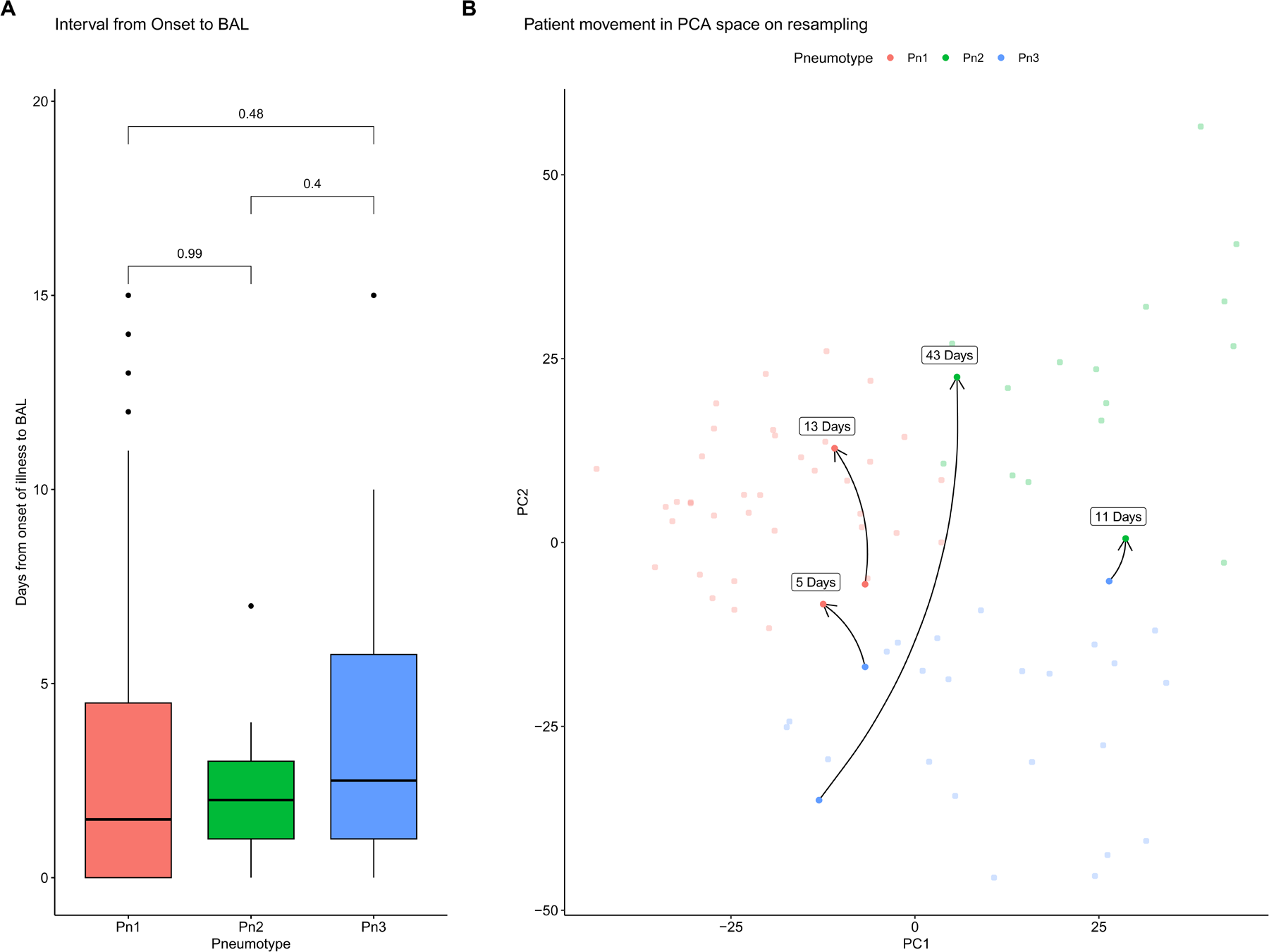
Relative indifference of pneumotype to temporal factors. **A** Boxplot of time interval from illness onset to time of BAL showing no significant differences between Pneumotypes. **B** Principal component analysis plot of the bronchoalveolar transcriptional clusters highlighting the four patients who were resampled. Arrows link the first and second resampling, with the gap in days between the samples indicated. For the patients resampled at 5, 11 and 13 days these were during the same admission and same episode of pneumonia, with resampling prompted by a deterioration in clinical picture and suspicion of recurrent infection. The patient sampled 43 days apart occurred on two separate ICU admissions with distinct episodes of suspected pneumonia

**Extended Data Figure 3.**
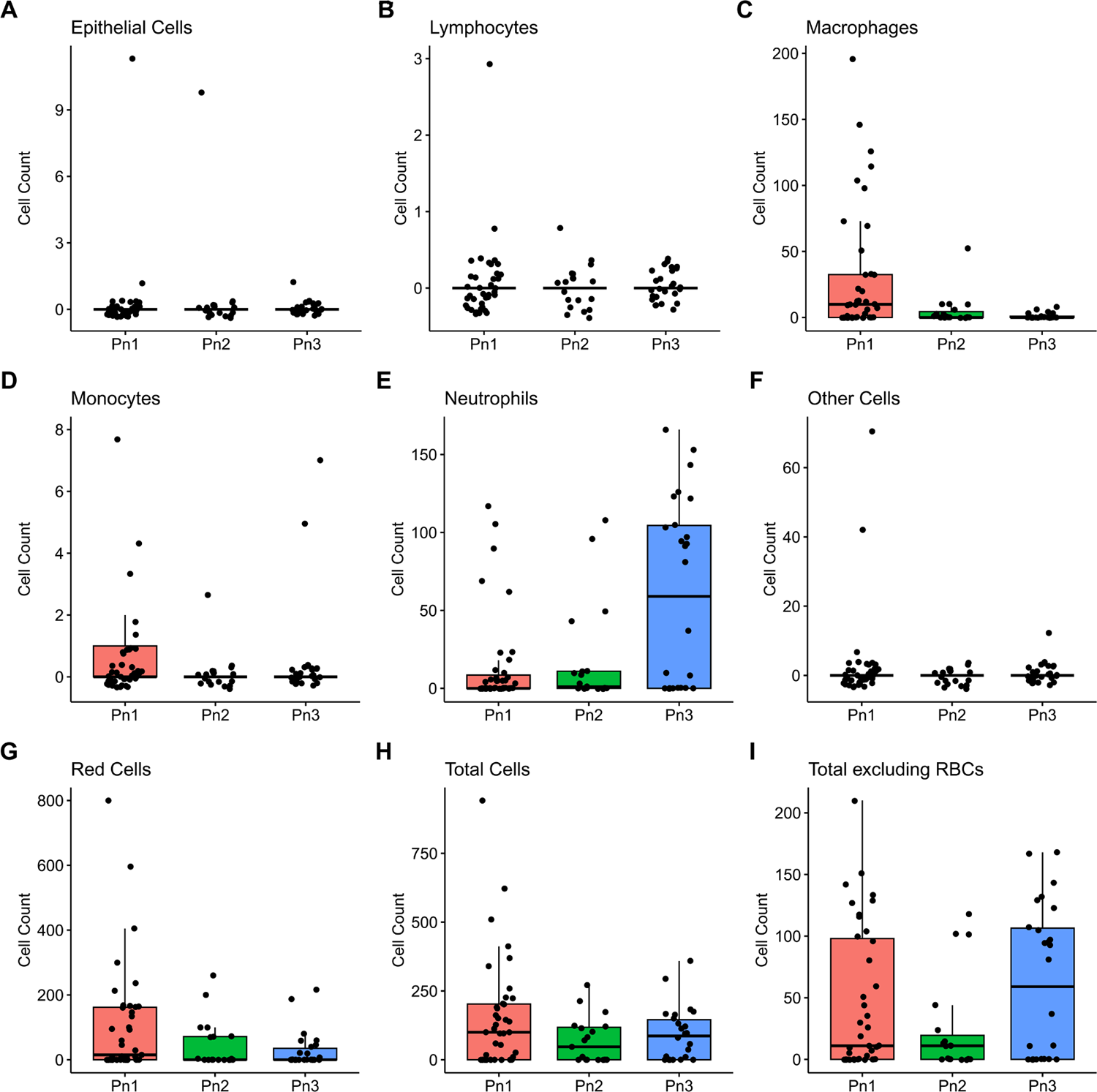
Differential cell counts by Cytospin followed by Kwik-Diff™ staining with identification by morphology, subdivided into Pneumotype. **A** Bronchial epithelial cells. **B** Lymphocytes. **C** Macrophages. **D** Monocytes. **E** Neutrophils. **F** Other cells. **G** Red blood cells. **H** Total cell count. **I** Total cell count excluding red blood cells.

**Extended Data Figure 4:**
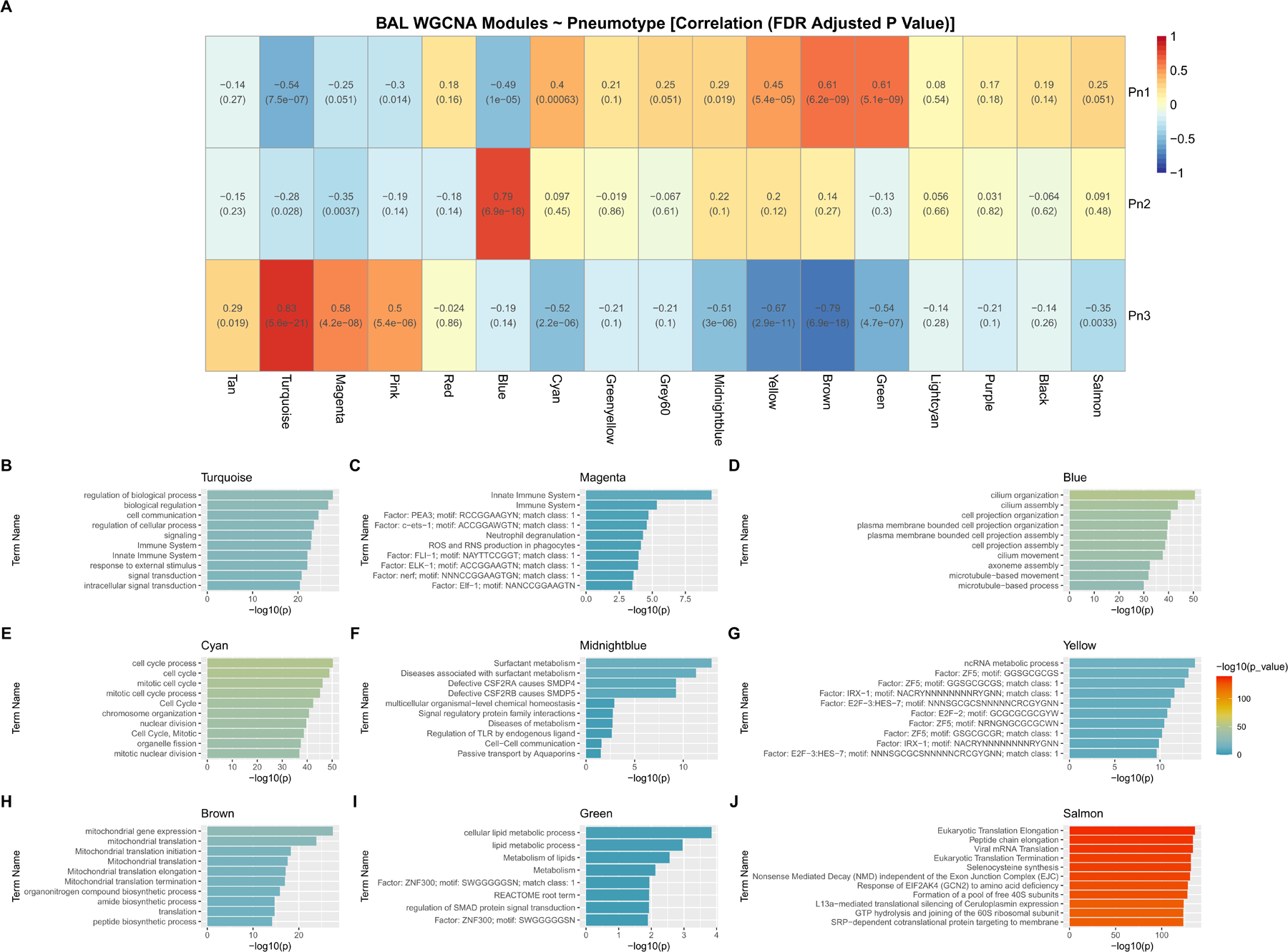
BAL Weight Gene Co-Expression Network Modules. **A** Heatmap of module correlations with each Pneumotype. **B-J** Over Representation Analysis of module hub genes for modules showing significant differences between Pneumotypes. Coloured by absolute −log10(p) to show differing magnitudes of significance between modules.

**Extended Data Figure 5:**
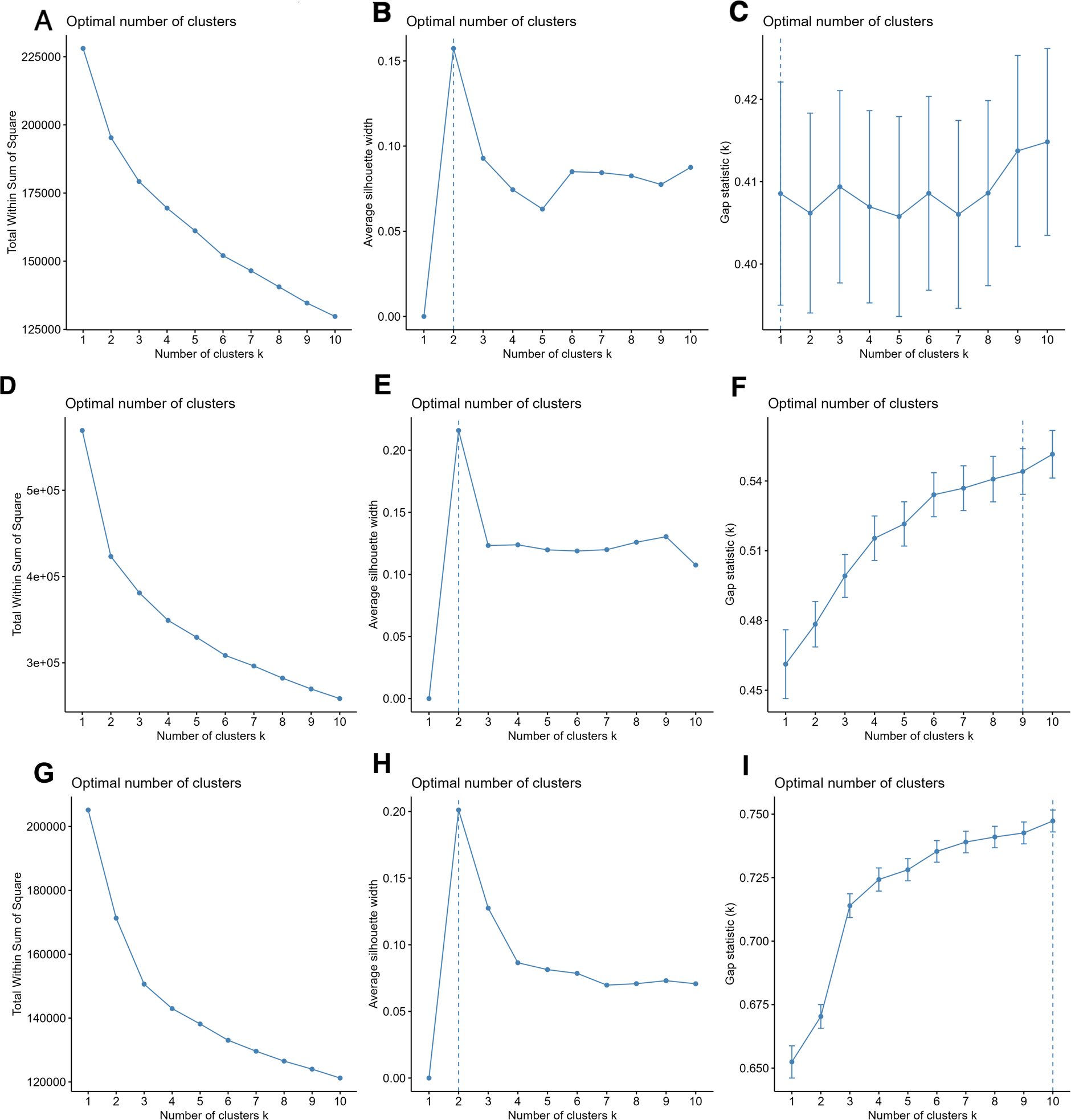
Clustering metrics for Blood clusters, tracheal aspirate bulk RNA sequencing data obtained from Langellier et al[19] and sorted alveolar macrophage RNA sequencing data obtained from Grant et al[43]. **A-C** Elbow plot, Silhouette Score and Gap statistic for blood bulk RNAseq samples. **D** Elbow plot of Langellier data demonstrating an elbow at 2 and no further reduction in with-group sum of squares between 2 and 5 clusters. **E** Silhouette score plot of Langellier data with maximal scores between one and two clusters. **F** Gap statistic of Langellier data plot indicating no clear local maximum. **G** Elbow plot of Grant data demonstrating an elbow at 3 and no further reduction in with-group sum of squares between 3 and 5 clusters. **H** Silhouette score plot of Grant data with maximal scores between two and three clusters. I Gap statistic plot of Grant data indicating a local maximum at 3.

**Extended Data Figure 6:**
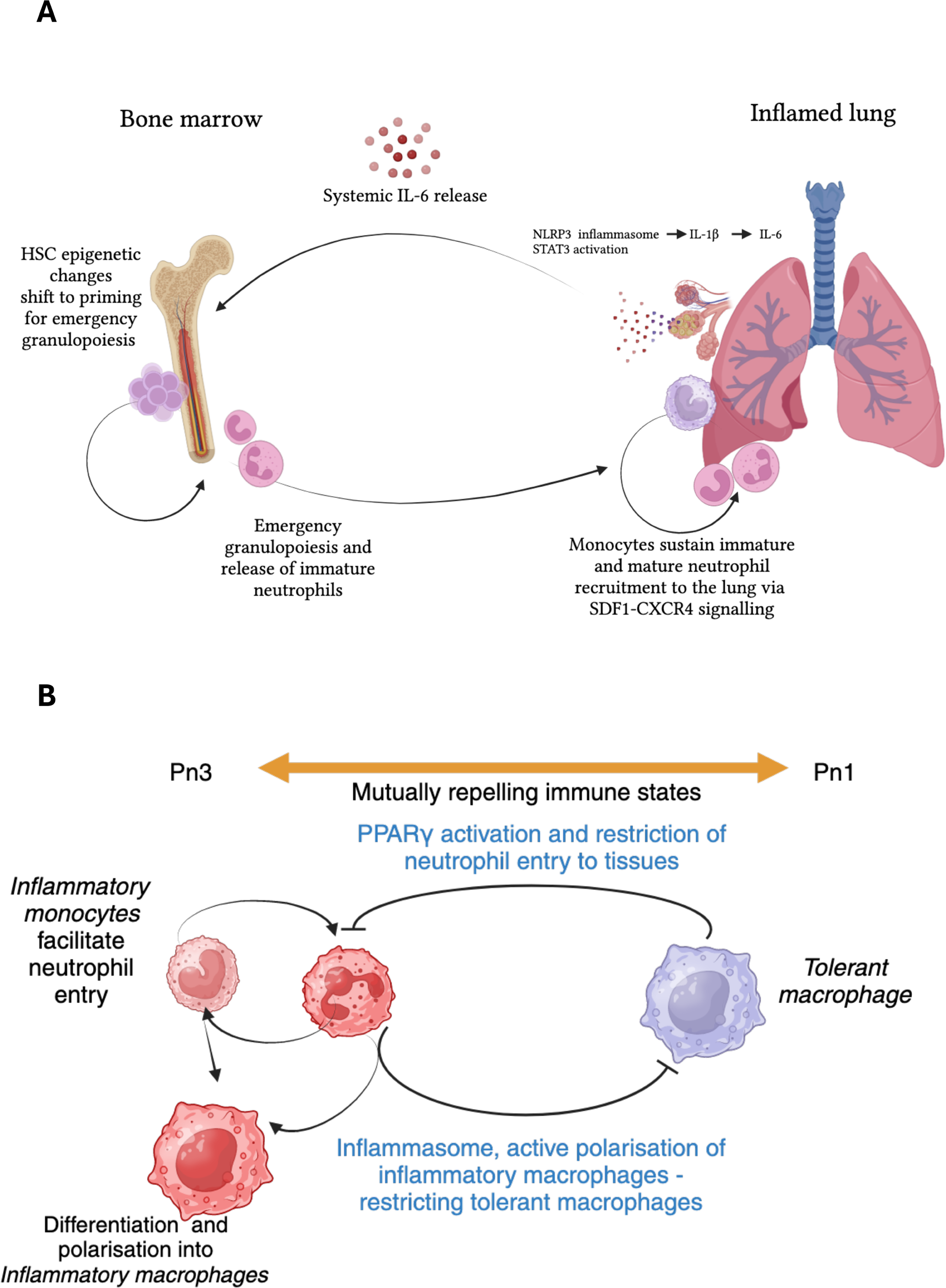
Schematic representations of hypothesised mechanisms underpinning Pneumotypes. **A** Schematic representation of the self-reinforcing and mutually reinforcing cycles that form a bi-stable equilibrium that drives sustained inflammation in Pn3, allowing inflammation to persist after removal of the precipitating insult. **B** Schematic representation of the mutually exclusive natures of Pn1 and 3, whereby either tolerant tissue-resident macrophages dominate and exclude neutrophils (Pn1) or neutrophils polarise macrophages to an inflammatory phenotype and produce a persistent peripheral blood infiltrative phenotype (Pn3).

## Methods

### Design/Setting/Participants

The study has been described previously[18]. Of the 95 patients recruited, sequenceable RNA that passed quality control was available from blood in 92 and BAL in 80 (5 patients were sampled twice with 4 having sequenceable RNA in the second sample), with both available for 79 patients.

Participants were recruited from a 20-bedded teaching hospital Intensive Care Unit (ICU). The unit is a mixed general medical-surgical unit which supports transplant and haematology-oncology services. Eligibility criteria were age ≥18, on mechanical ventilation, where the treating clinician suspected pneumonia and planned to undertake a diagnostic bronchoalveolar lavage (BAL). Exclusions were contraindications to bronchoscopy (e.g. by FiO2 >80%, severe hypercapnia, coagulopathy or presence of small diameter endotracheal tube) or lack of informed consent or proxy assent.

Eligible patients were included consecutively when the study team was available (the study team were routinely unavailable from Friday 5pm to Monday 8am, and also sporadically unavailable due to leave).

#### Ethical approvals

The prospective study was approved by the Leeds East Research Ethics Committee (17/YH/0286), Cambridge University Hospitals NHS Foundation Trust was the sponsor, and registered with clinicaltrials.gov (NCT03996330). The protocol has been deposited on Zenodo (doi/10.5281/zenodo.5081879). Written informed consent was obtained from patients or proxy assent, with retrospective consent sought from patients who regained capacity whilst in hospital. It was registered with Clinicaltrials.gov NCT03996330

#### Sampling

Patients underwent bronchoscopy and lavage as per the unit’s standard operating procedure. Following wedging of the scope in a radiologically affected subsegment, up to 200ml of saline were introduced in aliquots. The first aliquot of non-cellular material was discarded, and the remaining fluid processed for routine microbiology, Taqman array, microbial sequencing, host cell RNA sequencing, cytology and inflammatory protein assays. Simultaneous draw of arterial or venous blood from indwelling lines was collected into Paxgene tubes for RNA preservation (Preanalytix, Hombrechtikon, Switzerland). A further blood sample was collected into EDTA (Sarstedt, Nuembrecht, Germany) and used for plasma generation with downstream inflammatory protein assays.

### Isolation of host BAL cells

BAL was centrifuged at 700g for 5 minutes to pellet cells, cells were then resuspended in saline, counted and 2.5x 10^4^ cells were loaded in Shandon EZ Cytofunnels (ThermoFisher, Waltham, MA, USA) and spun onto cytoslides in a Cytospin cytocentrifuge at 32 × g (500 rpm) for 4 min. The slides were fixed in methanol for 5 minutes and cells were stained using Shandon Kwik-Diff stain (ThermoFisher) and counted manually. The remaining cells were centrifuged again, and the pellet was resuspended in 350ul RLT buffer (Qiagen) with b-mercaptoethanol and stored at –70oC until RNA extraction.

### RNA extraction and sequencing

RNA from BAL cells was extracted using an RNAeasy kit (Qiagen, Venlo, Netherlands), and RNA from blood stored in PAXgene tubes was extracted using a PAXgene Blood RNA Kit (PreAnalytiX), following manufacturer’s recommendations. Sequencing libraries were constructed using an NEB Ultra II RNA custom kit (New England Biolabs,Ipswich, MA, USA), cDNA was amplified with dual indexed tag barcodes (14 cycles) (Eurofins, Luxembourg), then purified using Agencourt AMPure XP SPRI beads (Beckman Coulter, Brea, CA, USA). Libraries were pooled in equimolar amounts (20-plex), normalised to 2.8nM and sequenced on the HiSeq 4000 platform (Ilumina, San Diego, CA, USA), to generate paired-end read lengths of 75 bp. Reads were mapped to the Genome Reference Consortium human build 38 (GRCh38) using Spliced Transcripts Alignment to a Reference (STAR) with read counts annotated using Ensembl 99. Following removal of the 12 haemoglobin genes from the blood samples, quality control checks were performed with FASTQc and Quality of RNA-Seq Toolset (QoRTs), resulting in the rejection of two lavage samples and two blood samples.

### Inflammatory protein analysis

Lavage supernatant and plasma inflammatory proteins were assayed using a Bio-Plex Pro Human Cytokine Screening 48-plex kit on a Bio-Plex 200 System (Bio-Rad, Hercules, CA, USA), following manufacturer’s recommendations.

### Microbiological assays

The microbiological processing for conventional culture, TaqMan array card (TAC) (ThermoFisher) and sequencing have been described in detail previously[18]. Briefly, samples were processed in accordance with the UK Standards for Microbiology Investigations (SMI) for conventional culture, alongside in-house PCR for respiratory viruses (adenovirus, enterovirus, human metapneumovirus, influenza A virus, influenza B virus, parainfluenza virus, rhinovirus, and respiratory syncytial virus), *Pneumocystis jirovecii* and herpesvirade (herpes simplex virus, human cytomegalovirus and Epstein Barr virus). The TaqMan array encompassed validated assays for 52 pathogens, with full details of coverage, development and validation with metagenomic sequencing reported previously[18].

### Adjudication of pneumonia

Diagnosis of pneumonia was independently assessed by 2 experienced clinicians with access to clinical, radiological and microbiological data who used pre-agreed criteria to independently rate cases as ‘definite’, ‘highly likely’, ‘unlikely’ or ‘not pneumonia’. Any disagreement was resolved by a 3rd clinician. ‘Confirmed pneumonia’ was defined as consensus of ‘definite’ or ‘highly likely’ in keeping with previous studies [58, 59]. Clinicians were blinded to host RNA and metagenomic sequencing results. The diagnostic components were summarized by assessing clinical, radiological, and microbiological criteria and the presence of systemic inflammation as defined by >=2 SIRS criteria (WCC <4 or >12, temp <36 or >38 degrees C, HR >90bpm, RR >20bpm). Clinical criteria were defined by an increase in frequency or volume of respiratory secretions, increased oxygen requirement, deterioration in compliance, or signs of pneumonia on clinical examination. Radiological criteria where new or worsening pulmonary infiltrates or consolidation on X-ray or Computed Tomography (CT) imaging not explained by another cause. Microbiological criteria were positive blood, sputum, or BAL culture for known respiratory pathogens, serological or urinary pneumococcal or legionella antigen, TAC detection with cycle time (CT) ≤32[18]. The patient’s location 48 hours prior to the onset of the illness being investigated was recorded as community or hospital, with subsequently confirmed pneumonias defined as community- or hospital-acquired.

### Clinical parameters

Baseline demographic information including age, sex, body mass index, comorbidities and primary reason for ICU admission was recorded. Admission APACHE 2 score and PaO2/FiO2 ratio, white cell count (WCC) and C-reactive protein (CRP) immediately prior to bronchoscopy were recorded.

Immunosuppression definition was based on the recent consensus statement[60] and consisted of neutropenia, hematological malignancy, HIV infection with detectable viral load/CD4 count <250, current administration of immunosuppressive medications including corticosteroids >20mg prednisolone equivalent and solid organ or bone marrow transplant. Patient outcomes were determined by electronic patient record (EPR) review after sufficient time for NHS spine updates to determine mortality up to 1 year and included duration of hospital admission and survival to nearest day, mechanical ventilation (end of last recorded period of mechanical ventilation) to nearest hour. Hazard ratios were calculated using Cox regression, with survival adjusted for age.

### Data management

Clinical data was collected from the EPR and recorded in a secure database. Patients were assigned unique study identifiers and identifiable information removed prior to analysis. Anonymized data and analysis code are made available with this publication [link].

### Potential sources of bias

Sources of bias with limited recourse for control were patient fitness for bronchoscopy and single-center recruitment. Although bronchoscopy forms part of the routine diagnostic workup for severe pneumonia in the trial unit, patients with difficult ventilation, on >80% oxygen or with significant coagulopathy will have been excluded by the treating clinicians.

To minimize batch effects during RNAseq, prepared libraries were stored frozen and sequenced as one batch. As this meant prolonged storage time for early samples, this was recorded. In addition, bronchoalveolar lavage has variable concentrations of RNA compared to blood. To assess the impact of this, return volume and whether lavage volume was <200ml was also recorded. Impact of technical and clinical co-variates was assessed by variance partitioning and principal component analysis, with final differential expression model controlling for age, sex, freezer time and return volume.

### Bioinformatics and statistics

#### RNAseq Quality Control

Batch effects and outliers checked for using Hierarchical clustering and scatterplots of non-zero genes by library size. Filtering was performed using the edgeR function “filterByExpr” which keeps genes with a Counts Per Million (CPM) >= minimum count divided by median library size multiplied by 1e6. After assessing CPM density plots pre and post-normalization, a filtering threshold minimum count of 20 in at least 10% of samples was set for BAL. Blood samples were less sparse and the default minimum count of 10 was appropriate. Variance stabilizing transformation[20] was applied prior to further analysis, with the exception of xCell deconvolution where TPM normalization was used on the advice of the package authors[22]. For 60,664 unique genes were sequenced, with 15,039 post-filtering in BAL and 14,620 in blood.

#### Clustering

The 10% most variable genes were used for clustering. Agglomerative, hybrid hierarchical k-means (HKMC) clustering using Euclidean distance and Ward’s method with 10 iterations of k-means consolidation was performed[21]. Three clusters in BAL and two in blood were identified based on elbow plots, a local maximum in the gap statistic (for BAL) and silhouette score (Extended Data Figures 1A-C and 4A-C). In the Langelier Tracheal Aspriate cohort[19] the elbow plot and silhouette score identify 2 clusters, with a continually increasing gap score suggesting these may not be well separated (Extended Data Figure 5D-F). The Grant sorted macrophage cohort[43] clearly identifies 3 clusters on elbow plot and gap statistic, though the maximum silhouette score was at 2 (Extended Data Figure 5G-I). HKMC was chosen over hierarchical and k-means clustering due to greater stability assessed by average pairwise Rand index on 100 bootstrapped samples with replacement[61]. Downstream clinical, microbiological and inflammatory protein features were robust to clustering method.

#### Deconvolution

Bulk RNA deconvolution to estimate cellularity was performed using xCell[22]. xCell performs cell type enrichment analysis for 64 cell signatures, pretrained on high-quality data. As recommended by package authors, TPM normalized gene counts were used for enrichment analysis, and spillover compensation utilized the default alpha=0.5. For comparison of BAL and TA samples, common genes raw counts were merged prior to TPM normalization - though without sample overlap these should be interpreted with caution as batch effects cannot be assessed.

#### Differential Expression

Differentially expressed (DE) genes between clusters were identified using an edgeR[62] and Limma[63] workflow. edgeR uses a negative binomial distribution and robust, quasi-likelihood dispersions were estimated after effective library size calculation.

Model design was informed by variance partitioning, principal component analysis and WGCNA module correlation with technical variables, the final model was 0 + cluster + age + sex + library storage time + BAL return volume. Library storage time was significantly correlated with WGCNA modules related to cell cycle, whilst BAL return volume explained 14% of variance in PC1, which was co-correlated with predicted macrophage proportion.

DE was tested relative to a log2 fold change threshold >1. The resulting p-value histogram for one vs rest was bimodal. This could not be rescued by more stringent gene filtering and is likely a product of DE testing on clusters, as these are defined by the variance of the dataset and inherently paired and complementary with respect to gene expression, reassuringly global significance testing using a quasi-likelihood test produced the desired anti-conservative pattern. Pi0 will therefore be inflated for one vs rest comparisons, resulting in overly conservative false discovery rate (FDR) correction and increased risk of type 2 error. FDR p-value threshold was set to 0.05.

#### Weighted gene co-expression network analysis (WGCNA)

WGCNA[64] analysis was performed on the BAL, blood and validation cohorts tracheal aspirate samples. The lowest soft-thresholding power that achieved a scale-free topology was used. In consensus module analysis of BAL and tracheal aspirate samples the recommended default of 12 was used as filtering thresholds had competing impacts on the optimal power. Signed networks were constructed in a single block with a minimum module size of 30 and dynamic tree cutting. Modules with a cut height <0.25 were merged. Module membership was calculated as the Pearson correlation between the normalized count and the module eigengene with FDR correction.

#### Pathway analysis

Pathway enrichment was performed on differentially expressed genes and WGCNA module hub genes (defined as absolute module membership >0.8) through g:Profiler R client[65]. For WGCNA modules, Over Representation Analysis (ORA) was performed against Gene Ontology Biological Processes (excluding “inferred from electronic annotation” evidence codes), Reactome and TRANSFAC databases using a custom background with a significance threshold <0.05 after g:SCS correction.

Unlike Benjamini-Hochberg FDR correction, the g:SCS algorithm does not assume test independence, an assumption necessarily violated by hierarchical gene ontology terms. Geneset Enrichment Analysis (GSEA) was performed on differentially expressed genes using ClusterProfiler R package ranked by log-fold change, with Benjamini-Hochberg correction (g:SCS not available for GSEA) and a significance threshold of <0.05. Network plots were constructed from significantly enriched pathways and their core enriched genes (or overlapping genes for ORA). Protein-Protein Interactions between core enriched genes with >80% confidence from StringDB (strind-db.org, version 12) were incorporated into the network as edges.

Upstream transcription factor prediction was performed using Transcription Factor Enrichment Analysis using ChEA3[23]. This analysis was conducted separately for positively and negatively differentially expressed genes for each Pneumotype as differential expression on clusters produces complementary gene lists – with each upregulated gene appearing downregulated in another cluster.

#### Survival and time to extubation analysis

Survival was assessed with censoring at 1 year using a Cox proportional hazards regression model including Pneumotype and age (HR 1.03, p=0.036). Time to extubation was assessed using Pneumotype alone as age had no effect in this model (HR 1.00, p=0.7). Severity of illness and respiratory failure were not included as it is hypothesized that Pneumotype may have causal influence on these. Kaplan-Meier curves where also fit (Figure 1F-G). Analysis was performed using the survival R package.

#### Inflammatory proteins

Measured inflammatory proteins (pg/ml) were not normally distributed and showed significant skew. Differences between Pneumotypes were assessed using Kruskal Wallis Rank Sum and P values were FDR adjusted for 96 comparisons (48 BAL and 48 plasma). Pearson Correlation Coefficients were calculated on log1p transformed values and P values calculated using Student’s asymptotic p-value for correlation then FDR adjusted for 48×48 comparisons. Following log1p transformation skewness was not <|2| for plasma SDF-1a, IL-9, MIP-1a and BAL MIF, GM-CSF and IL-13 and correlation should be interpreted with caution for these proteins. Scatter plots for highly correlated inflammatory proteins were inspected to assess the influence of outliers.

#### Validation

No equivalent cohort of suspected adult pneumonia patients with whole BAL RNAseq was identified, however Langelier *et al.*[19] have made tracheal aspirate RNAseq and clinical data available for a closely related clinical cohort of ventilated patients being investigated for suspected pneumonia. Whilst Grant et al.[43] included RNAseq of flow-sorted macrophages and flow cytometry from BAL in patients with microbiologically confirmed pneumonia (and non-pneumonia ICU controls) as a comparator to COVID pneumonia.

Comparisons of tracheal aspirate and bronchoalveolar lavage host transcriptomes is not well described in the literature, and the impact of differing sampling methods is unknown. However, we attempted to replicate our clustering findings of enrichment for bacterial organisms and immunosuppression. Significance testing of these proportions was tested by Pearson’s Chi-squared test. WGCNA consensus module analysis was used to compare gene modules common to the BAL and TA datasets and thus similarities and differences in expression between clusters and datasets. As tracheal aspirates are less likely to sample alveolar cells, this effect was estimated using deconvolution on the pooled BAL and TA data.

In the Grant cohort bulk RNAseq was limited to flow-sorted macrophages and thus any epithelial cell transcription defining Pn2 could not be assessed. However, this data clustered on M1/M2 macrophage polarization, and the flow cytometry data for these samples could be used to validate the predicted cellularity associated with this in Pn1 and Pn3.

